# Spatial Profiling of Lung SARS-CoV-2 and Influenza Virus Infection Dissects Virus-Specific Host Responses and Gene Signatures

**DOI:** 10.1101/2020.11.04.20225557

**Authors:** Arutha Kulasinghe, Chin Wee Tan, Anna Flavia Ribeiro dos Santos Miggiolaro, James Monkman, Dharmesh Bhuva, Jarbas da Silva Motta Junior, Caroline Busatta Vaz de Paula, Seigo Nagashima, Cristina Pellegrino Baena, Paulo Souza-Fonseca-Guimaraes, Lucia de Noronha, Timothy McCulloch, Gustavo Rodrigues Rossi, Caroline Cooper, Benjamin Tang, Kirsty R. Short, Melissa J Davis, Fernando Souza-Fonseca-Guimaraes, Gabrielle T. Belz, Ken O’Byrne

## Abstract

The severe acute respiratory syndrome coronavirus 2 (SARS-CoV-2) that emerged in late 2019 has spread globally, causing a pandemic of respiratory illness designated coronavirus disease 2019 (COVID-19). Robust blood biomarkers that reflect tissue damage are urgently needed to better stratify and triage infected patients. Here, we use spatial transcriptomics to generate an in-depth picture of the pulmonary transcriptional landscape of COVID-19 (10 patients), pandemic H1N1 (pH1N1) influenza (5) and uninfected control patients (4). Host transcriptomics showed a significant upregulation of genes associated with inflammation, type I interferon production, coagulation and angiogenesis in the lungs of COVID-19 patients compared to non-infected controls. SARS-CoV-2 was non-uniformly distributed in lungs with few areas of high viral load and these were largely only associated with an increased type I interferon response. A very limited number of genes were differentially expressed between the lungs of influenza and COVID-19 patients. Specific interferon-associated genes (including *IFI27*) were identified as candidate novel biomarkers for COVID-19 differentiating this COVID-19 from influenza. Collectively, these data demonstrate that spatial transcriptomics is a powerful tool to identify novel gene signatures within tissues, offering new insights into the pathogenesis of SARS-COV-2 to aid in patient triage and treatment.

## Introduction

Since its emergence in 2019, severe acute respiratory syndrome-associated coronavirus-2 (SARS-CoV-2) has caused a broad spectrum of disease, ranging from asymptomatic infections to acute respiratory distress syndrome (ARDS). Despite a lower fatality rate than other related coronaviruses such as SARS-CoV-1 and middle east respiratory syndrome coronavirus (MERS-CoV), SARS-CoV-2 is highly transmissible and to date has resulted in over >45M infections and >1.19M deaths worldwide and rising^1^. COVID-19 is the clinical manifestation of SARS-CoV-2 infection.

ARDS develops in 42% of COVID-19 patients presenting with pneumonia and accounts for a significant number of deaths associated with COVID-19^2,3^. ARDS is a form of hypoxemic respiratory failure defined by the presence of diffuse alveolar damage (DAD) commonly associated with bacterial pneumonia, sepsis, pancreatitis or trauma. The pathogenesis of ARDS is typically attributed to inflammatory injury to the alveolar-capillary membrane which leads to pulmonary oedema and respiratory insufficiency. In response to injury, alveolar macrophages orchestrate inflammation of the lung by recruiting neutrophils and circulating macrophages to the site of injury leading to the death of alveolar epithelial cells and further impairing lung function^4^. Severe COVID-19 is often also characterised by concurrent vascular disease and coagulopathy. This includes breakdown of the vascular barrier, oedema, endothelialitis and thrombosis. Thrombotic and microvascular complications are frequently recorded in deceased patients, suggesting that vascular pathology is a major driver of severe disease^5^. While these mechanisms contribute to disease progression, it is unclear which mechanisms are the predominant factors in causing fatality. Biomarkers can be used to delineate these separate underlying mechanisms thereby clarifying the association between disease mechanism and fatality. Once validated, these biomarkers can also help stratify clinical care for patients and guide management of those with severe disease.

Transcriptomic analysis of patient tissue offers a unique opportunity to understand the pathogenesis of SARS-CoV-2, to define its short and long-term complications and to identify reliable biomarkers to help triage patients. To date, the major search for clinical biomarkers, including transcriptomic analyses on COVID-19 patient samples, have been performed on whole blood. Blood samples from infected patients are highly accessible but it is not clear whether blood parameters accurately reflect either the viral load or the level of tissue damage that clinical treatments aim to control^6^. The lung is the primary site of viral replication, yet only a limited number of transcriptomic studies have been performed on the lungs of COVID-19 patients. Such studies have used transcriptomic analyses on only a small patient cohort (1-2 patients)^7,8^ or relied on ‘bulk’ sequencing of whole lung tissue, where any insight into tissue heterogeneity in this complex organ is lost ^5,9,10^. In contrast, gene expression panels applied directly on tissue sections take into account the spatial location of transcriptomic features and are able to directly sample intra-organ heterogeneity. This provides a much deeper picture of the cellular changes driven by viral infection, and is a powerful way to define the characteristics of the host response to the virus and the spatial relationships between them. Indeed, preliminary spatial transcriptomic analysis of COVID-19 patient lungs appears promising^11^. However, such analyses require the application of advanced bioinformatics techniques to control for the numerous potential confounders present in gene expression data sampled in such a complex experimental design. In addition, there is a clear need to distinguish the host SARS-CoV-2 transcriptomic response (and associated spatial heterogeneity) from other infectious triggers of ARDS such as influenza virus, which may be circulating concurrently with SARS-CoV-2. Bulk sequencing data already suggests that these two infections can be distinguished by the strength of the thrombotic and coagulant response in the lungs of COVID-19 patients^5^. This differential analysis becomes even more pertinent as the Northern Hemisphere enters their annual influenza season and there may be increased demand for biomarkers that can rapidly differentiate influenza from COVID-19.

To address these questions, we investigated the pathogenesis of SARS-CoV-2 in lung tissue using spatial transcriptomics approaches from rapid autopsy samples taken from 10 COVID-19 patients, five 2009 pandemic H1N1 (pH1N1) influenza virus patients and four control patient samples collected at a single institution. GeoMX^™^ Digital Spatial Profiling (DSP) combined with bioinformatic modelling allowed deconvolution of individual variability from viral and host factors. These data showed that viral load has a heterogenous impact within a patient on the pulmonary transcriptomic response there is significant overlap in the gene expression profiles between influenza virus and SARS-CoV-2 infected samples. However, several unique transcriptomic signatures were detected in the lungs of COVID-19 patients. This study demonstrates the strengths of spatial transcriptomics coupled with powerful linear modelling bioinformatics modelling techniques to broadly identify key pathways involved in viral infections and to clearly differentiate the involvement of distinct cellular and molecular pathways in different infections.

## Results

### Patient Characteristics

Lung tissues were obtained by rapid autopsy from 10 SARS-CoV-2 infected patients, five pH1N1 influenza virus infected patients and four non-virally infected control patients. The SARS-CoV-2 cohort was made up of four females and six males with mean age of 76 years (ranging from 46-93 years). These patients exhibited multiple comorbidities including obesity, diabetes and heart disease and had received varying treatment following hospital admission. Patient survival after admission ranged from 8 to 38 days, with time on mechanical ventilation ranging from 3 to 21 days (Extended Data Table 1). Blood profiling performed on admission and again immediately prior to death showed elevated D-dimer levels for 7 of 10 patients consistent with vascular coagulopathy (Extended Data Table 2).

**Table 1.** Geneset Enrichment Analysis for SARS-CoV-2 vs uninfected DE genes using Molecular Signatures Database (MSigDB) Hallmark gene sets.

| Geneset Name <sup>a,b</sup> | N Genes <sup>c</sup> | Direction <sup>d</sup> | two-sided directional |  | non-directional |  |
| --- | --- | --- | --- | --- | --- | --- |
|  |  |  | p-value <sup>e</sup> | FDR <sup>f</sup> | p-value <sup>e</sup> | FDR <sup>f</sup> |
| HALLMARK_REACTIVE_OXYGEN_SPECIES_PATHWAY | 13 | Up | 2.39E-09 | 1.19E-07 | 1.83E-03 | 2.47E-03 |
| HALLMARK_COMPLEMENT | 76 | Up | 1.45E-08 | 2.54E-07 | 3.59E-24 | 1.80E-22 |
| HALLMARK_KRAS_SIGNALING_DN | 40 | Down | 1.52E-08 | 2.54E-07 | 9.34E-12 | 2.12E-11 |
| HALLMARK_IL6_JAK_STAT3_SIGNALING | 73 | Up | 2.76E-08 | 3.45E-07 | 1.93E-20 | 2.41E-19 |
| HALLMARK_UNFOLDED_PROTEIN_RESPONSE | 16 | Up | 4.85E-08 | 4.27E-07 | 3.40E-03 | 4.47E-03 |
| HALLMARK_APOPTOSIS | 79 | Up | 5.12E-08 | 4.27E-07 | 1.54E-18 | 9.62E-18 |
| HALLMARK_OXIDATIVE_PHOSPHORYLATION | 33 | Up | 7.87E-07 | 5.62E-06 | 5.30E-06 | 8.84E-06 |
| HALLMARK_P53_PATHWAY | 56 | Up | 1.69E-06 | 1.05E-05 | 3.64E-17 | 2.02E-16 |
| HALLMARK_MTORC1_SIGNALING | 47 | Up | 2.82E-06 | 1.45E-05 | 5.39E-13 | 1.42E-12 |
| HALLMARK_INTERFERON_GAMMA_RESPONSE | 110 | Up | 2.91E-06 | 1.45E-05 | 1.68E-14 | 5.59E-14 |
| HALLMARK_PROTEIN_SECRETION | 10 | Up | 3.66E-06 | 1.66E-05 | 7.27E-02 | 7.73E-02 |
| HALLMARK_GLYCOLYSIS | 48 | Up | 4.22E-06 | 1.76E-05 | 1.00E-14 | 3.59E-14 |
| HALLMARK_PANCREAS_BETA_CELLS | 5 | Down | 6.76E-06 | 2.60E-05 | 8.09E-01 | 8.09E-01 |
| HALLMARK_HYPOXIA | 60 | Up | 7.99E-06 | 2.85E-05 | 1.44E-19 | 1.44E-18 |
| HALLMARK_MYC_TARGETS_V2 | 8 | Up | 9.64E-06 | 3.21E-05 | 5.57E-01 | 5.69E-01 |
| HALLMARK_CHOLESTEROL_HOMEOSTASIS | 15 | Up | 1.51E-05 | 4.64E-05 | 1.47E-02 | 1.71E-02 |
| HALLMARK_INTERFERON_ALPHA_RESPONSE | 52 | Up | 1.58E-05 | 4.64E-05 | 1.84E-09 | 3.92E-09 |
| HALLMARK_SPERMATOGENESIS | 19 | Down | 7.45E-05 | 2.07E-04 | 2.59E-02 | 2.88E-02 |
| HALLMARK_MYC_TARGETS_V1 | 28 | Up | 9.53E-05 | 2.51E-04 | 3.99E-05 | 6.04E-05 |
| HALLMARK_KRAS_SIGNALING_UP | 77 | Up | 1.68E-04 | 4.19E-04 | 7.44E-17 | 3.72E-16 |
| HALLMARK_ADIPOGENESIS | 30 | Up | 2.11E-04 | 5.03E-04 | 1.32E-05 | 2.13E-05 |
| HALLMARK_COAGULATION | 56 | Up | 3.95E-04 | 8.97E-04 | 7.17E-16 | 3.26E-15 |
| HALLMARK_XENOBIOTIC_METABOLISM | 36 | Up | 4.81E-04 | 1.04E-03 | 2.36E-12 | 5.63E-12 |
| HALLMARK_ANDROGEN_RESPONSE | 16 | Up | 6.64E-04 | 1.38E-03 | 5.69E-03 | 7.30E-03 |
| HALLMARK_FATTY_ACID_METABOLISM | 20 | Up | 8.12E-04 | 1.62E-03 | 2.32E-05 | 3.63E-05 |
| HALLMARK_APICAL_JUNCTION | 61 | Up | 1.05E-03 | 2.03E-03 | 2.65E-19 | 2.01E-18 |
| HALLMARK_UV_RESPONSE_UP | 46 | Up | 1.12E-03 | 2.07E-03 | 8.56E-13 | 2.14E-12 |
| HALLMARK_EPITHELIAL_MESENCHYMAL_TRANSITION | 81 | Up | 1.28E-03 | 2.29E-03 | 1.15E-15 | 4.78E-15 |
| HALLMARK_ESTROGEN_RESPONSE_EARLY | 40 | Up | 1.46E-03 | 2.52E-03 | 3.49E-09 | 6.98E-09 |
| HALLMARK_TGF_BETA_SIGNALING | 20 | Up | 1.78E-03 | 2.97E-03 | 4.42E-04 | 6.32E-04 |
| HALLMARK_ESTROGEN_RESPONSE_LATE | 51 | Up | 1.92E-03 | 3.10E-03 | 3.36E-13 | 9.88E-13 |
| HALLMARK_DNA_REPAIR | 27 | Up | 6.58E-03 | 1.03E-02 | 1.10E-06 | 1.96E-06 |
| HALLMARK_MITOTIC_SPINDLE | 27 | Up | 7.55E-03 | 1.14E-02 | 7.56E-04 | 1.05E-03 |
| HALLMARK_IL2_STAT5_SIGNALING | 85 | Up | 1.09E-02 | 1.56E-02 | 2.08E-15 | 7.99E-15 |
| HALLMARK_ANGIOGENESIS | 19 | Up | 1.09E-02 | 1.56E-02 | 5.94E-03 | 7.42E-03 |
| HALLMARK_UV_RESPONSE_DN | 44 | Up | 1.20E-02 | 1.66E-02 | 1.88E-09 | 3.92E-09 |
| HALLMARK_HEDGEHOG_SIGNALING | 12 | Up | 1.32E-02 | 1.78E-02 | 1.05E-01 | 1.09E-01 |
| HALLMARK_G2M_CHECKPOINT | 67 | Up | 1.51E-02 | 1.99E-02 | 3.69E-13 | 1.02E-12 |
| HALLMARK_APICAL_SURFACE | 13 | Up | 1.62E-02 | 2.07E-02 | 4.80E-02 | 5.22E-02 |
| HALLMARK_TNFA_SIGNALING_VIA_NFKB | 107 | Up | 2.16E-02 | 2.70E-02 | 6.21E-14 | 1.94E-13 |
| HALLMARK_INFLAMMATORY_RESPONSE | 115 | Up | 2.64E-02 | 3.22E-02 | 3.08E-21 | 5.14E-20 |
| HALLMARK_E2F_TARGETS | 61 | Up | 2.91E-02 | 3.47E-02 | 9.03E-09 | 1.74E-08 |
| HALLMARK_NOTCH_SIGNALING | 21 | Up | 4.76E-02 | 5.53E-02 | 1.95E-04 | 2.87E-04 |
| HALLMARK_MYOGENESIS | 35 | Up | 6.89E-02 | 7.76E-02 | 4.24E-08 | 7.85E-08 |
| HALLMARK_PI3K_AKT_MTOR_SIGNALING | 56 | Up | 6.98E-02 | 7.76E-02 | 2.81E-19 | 2.01E-18 |
| HALLMARK_PEROXISOME | 17 | Up | 1.46E-01 | 1.58E-01 | 7.60E-03 | 9.05E-03 |
| HALLMARK_ALLOGRAFT_REJECTION | 141 | Up | 3.56E-01 | 3.79E-01 | 4.75E-23 | 1.19E-21 |
| HALLMARK_WNT_BETA_CATENIN_SIGNALING | 27 | Up | 4.04E-01 | 4.21E-01 | 2.65E-06 | 4.57E-06 |
| HALLMARK_BILE_ACID_METABOLISM | 14 | Down | 5.53E-01 | 5.64E-01 | 1.63E-02 | 1.85E-02 |
| HALLMARK_HEME_METABOLISM | 19 | Up | 9.10E-01 | 9.10E-01 | 6.52E-03 | 7.95E-03 |

<sup>a</sup> Geneset signatures in green are enriched in SARS-CoV-2 compared to control (i.e. Direction is Up) while red is the opposite direction (i.e. Down). <sup>b</sup> Genesets highlighted in yellow indicates biological processes upregulated in SARS-CoV-2 infected lungs. List ordered by descending FDR. <sup>c</sup> N-Genes, number of genes in the geneset. <sup>d</sup> Direction, direction of change (i.e. up or down). <sup>e</sup> p-value, statistical significance of the difference. <sup>f</sup> FDR, false discovery rate.

**Table 2.**
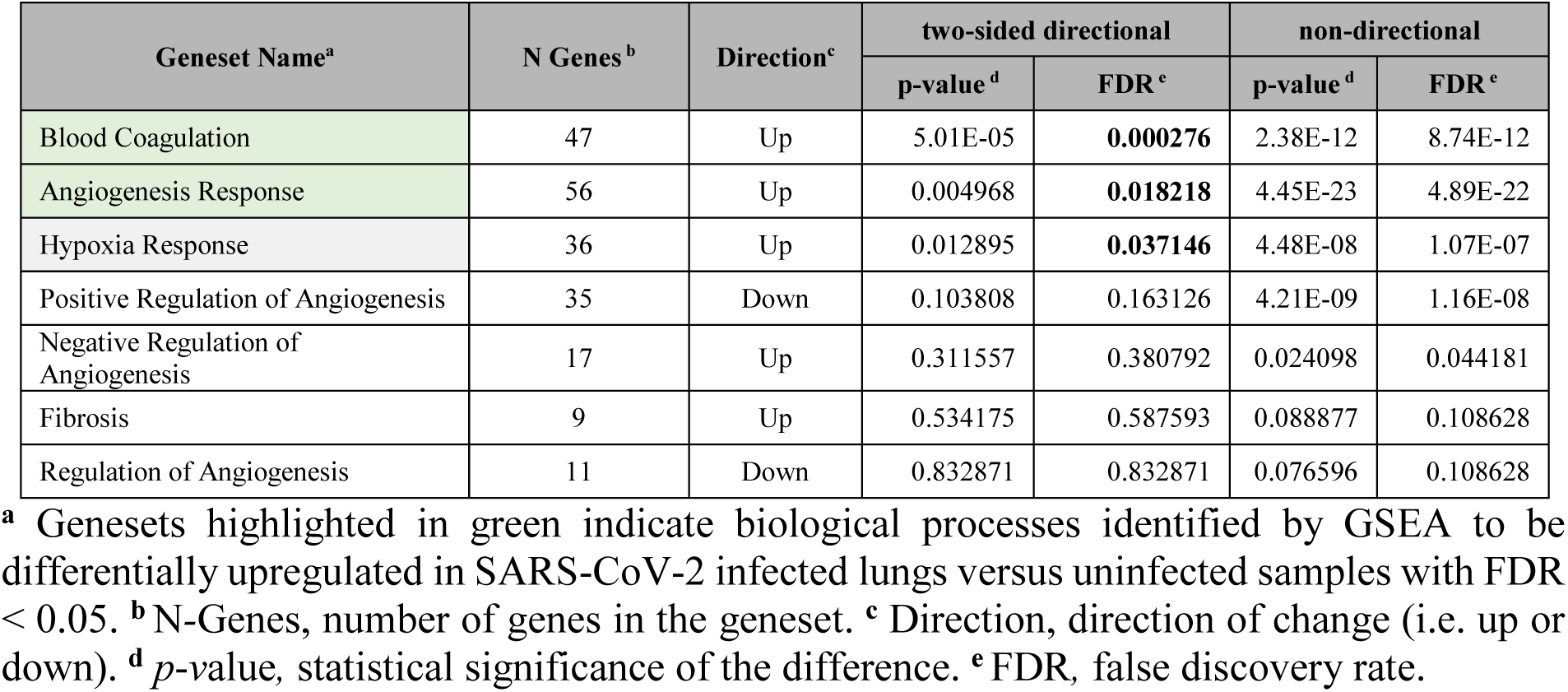
Geneset Enrichment Analysis for SARS-CoV-2 vs uninfected DE genes using nanoString’s nCounter® PanCancer Progression Panel custom gene sets.

The pH1N1 influenza virus patient cohort comprised five males with a mean age of 45.8 years (ranging from 31-53 years). All patients had received mechanical ventilation from the time of admission until death. The uninfected control cohort consisted of four males with a mean age of 36.25 (1 ranging from 18-60 years) whose tissues were donated following death from non-viral and non-respiratory diseases (Extended Data Table 3).

**Table 3.** Genes differentially expressed in the lungs of SARS-CoV-2 patients with high viral load compared with patients with low viral load (patient-based analysis).

| Gene name <sup>a</sup> | Fold Change (log2) | Average Expression | p-value <sup>b</sup> | Adjusted p-value <sup>c</sup> |
| --- | --- | --- | --- | --- |
| *S* | 1.916 | 8.900 | 2.89E-05 | 9.82E-03 |
| *ORF1ab* | 1.861 | 9.036 | 3.88E-05 | 9.82E-03 |
| *CXCL10* | 1.761 | 9.153 | 9.32E-06 | 5.76E-03 |
| *IFIT3* | 1.346 | 9.616 | 1.41E-05 | 6.52E-03 |
| *ISG15* | 1.286 | 9.454 | 8.13E-05 | 1.67E-02 |
| *MX1* | 1.249 | 9.748 | 3.57E-06 | 3.31E-03 |
| *GBP1* | 1.151 | 9.005 | 1.33E-09 | 2.46E-06 |
| *IFI6* | 1.037 | 9.913 | 3.58E-04 | 3.72E-02 |
| *STAT1* | 0.927 | 9.820 | 4.24E-05 | 9.82E-03 |
| *IDO1* | 0.925 | 8.539 | 1.85E-04 | 2.64E-02 |
| *OAS3* | 0.795 | 8.783 | 3.51E-05 | 9.82E-03 |
| *RSAD2* | 0.792 | 8.662 | 5.23E-04 | 4.40E-02 |
| *TAP1* | 0.748 | 10.082 | 4.60E-04 | 4.05E-02 |
| *PSMB9* | 0.725 | 9.919 | 1.66E-04 | 2.55E-02 |
| *GBP4* | 0.677 | 9.248 | 1.44E-04 | 2.43E-02 |
| *HERC6* | 0.627 | 8.451 | 6.30E-04 | 4.87E-02 |
| *IFI35* | 0.485 | 9.026 | 4.57E-04 | 4.05E-02 |
| *NR3C1* | -0.313 | 8.839 | 3.62E-04 | 3.72E-02 |
| *ATF2* | -0.330 | 8.348 | 3.03E-04 | 3.50E-02 |
| *SERINC3* | -0.415 | 9.130 | 5.81E-04 | 4.68E-02 |
| *JAK1* | -0.479 | 9.550 | 1.37E-04 | 2.43E-02 |
| *FYN* | -0.518 | 8.918 | 2.50E-04 | 3.09E-02 |
| *C4BPA* | -0.728 | 8.366 | 2.44E-04 | 3.09E-02 |
| *SERPINA1* | -1.251 | 10.750 | 3.90E-04 | 3.80E-02 |

<sup>a</sup> Up-regulated genes in high viral load compared to low viral load SARS-CoV-2 lung samples are shown in green, downregulated genes shown in red. <sup>b</sup> *p*-value, statistical significance of the difference. <sup>c</sup> *adjusted p*-value, the smallest familywise significance level for multiple comparison testing

### Histopathological Characterisation

Tissue microarrays (TMAs) were prepared from pulmonary formalin-fixed paraffin-embedded cores of each of the three cohorts (one core per patient) and blinded histological examination performed by a pathologist as shown in Extended Data Table 4. Acute phase diffuse alveolar damage (DAD) characterised by hyaline membranes, type 2 pneumocyte hyperplasia and alveolar septal thickening was a prominent feature of SARS-CoV-2 cores. pH1N1 tissues also showed features of DAD, with alveolar haemorrhage evident in two of the cores. These findings were not seen in control cores. Fibrosis was not a prominent feature apart from one SARS-CoV-2 core that exhibited moderate interstitial and alveolar fibrosis with concordant histological organising pneumonia. Two pH1N1 cores showed mild interstitial fibrosis that were characterised by acute phase or late organising phase DAD.

### RNAscope^®^ and ROI Selection for Digital Spatial Transcriptional Profiling

Transcriptional profiling of target cores was guided by the detection of SARS-CoV-2 spike mRNA. Two cores (LN1 and LN3) exhibited strong signals for viral mRNA (Fig. 1a) and these cores were comprehensively evaluated using the GeoMx DSP platform (shaded regions, Fig. 1b). Twenty-five regions of interest (ROIs) were selected from these two RNAscope® positive cores and 21 ROIs were selected from remaining eight cores of SARS-CoV-2 infected patients (minimum of one ROI per patient) for which viral mRNA was below detection by RNAscope. Eight ROIs were selected from the lungs of five pH1N1 patients (minimum of one ROI per patient), and five ROIs were selected from the lungs of control patients (minimum of one ROI per patient).

**Figure 1.**
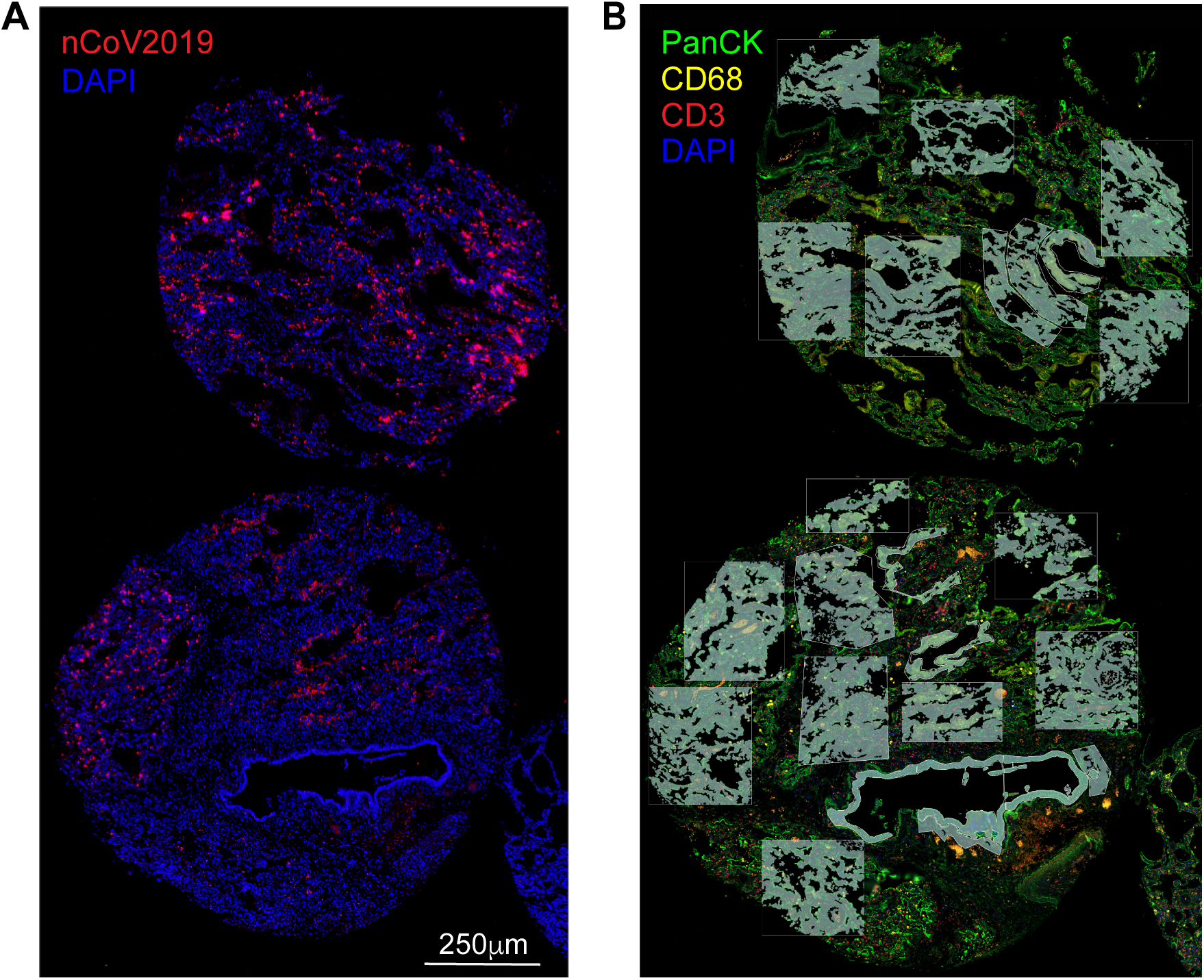
RNA-FISH and digital spatial profiling morphology marker visualisation of nCoV2019 mRNA positive cores. ROI selection was guided by RNAscope-FISH staining for nCoV2019 spike mRNA. **A**. RNA-FISH staining on COVID TMA showing the cores LN1 and LN3 which were highly positive for nCoV2019 spike mRNA (red). Nuclei are shown in blue (DAPI staining). Scale bar, 1000 µm. B. Representative ROIs selected across LN1 and LN3 SARS-CoV-2 virus positive cores on immunohistological paraffin embedded sections stained for monocytes (CD68, yellow), T cells (CD3ε, red), lung epithelial cells (PanCK, green) and nuclei (DAPI, blue). Scale bar, 500 µm.

SARS-CoV-2 RNAscope^®^ abundance was semi-quantitatively assessed for each ROI. Representative scoring criteria from 0 to 3 are shown in Extended Data Fig. 1 where positive scores were allocated only to regions within LN1 and LN3 (Extended Data Table 5). In addition, concordant spike protein immunohistochemistry (IHC) appeared to be specific for type 2 pneumocytes and bronchiolar epithelial regions, however, some staining was also observed within interstitial lymphocytes and alveolar macrophages (Extended Data Fig. 1). Interestingly, spike protein IHC indicated virus in cores other than LN1 and LN3 (Extended Data Table 5) that did not produce a detectable signal using RNAscope^®^. Low level non-specific staining was observed in some regions of pH1N1 and control tissues Extended Data Table 5. We thus focussed our ROI selection on cores that indicated high mRNA integrity and viral detection by RNAscope^®^.

### Histopathology and Dominant Features of ROIs

Assessment of histology by a pathologist allowed ROIs to be grouped by tissue architecture prior to further analysis. The predominant cell types found within each ROI were annotated in addition to scoring of their histopathology and spike protein IHC (Extended Data Table 5). ROIs could be broadly categorised into regions dominated by bronchiolar epithelial structures, type 2 pneumocytes, hyaline membranes and macrophages (Extended Data Fig. 2). Alveolar oedema, acute inflammation and squamous metaplasia were not observed in diseased or control cores, however, type 2 pneumocyte hyperplasia and capillary congestion were present in SARS-CoV-2 pulmonary tissues.

### Data normalization strategy and exploratory analysis of spatial transcriptomic data

The transcriptomic profile of the lungs of deceased patients was then examined using spatial transcriptomics. Data was corrected for systematic bias to allow for comparisons between SARS-CoV-2, pH1N1 influenza virus and control samples (Extended Data Fig. 3). Principal components analysis (PCA) plots were then used to explore the variability in SARS-CoV-2, pH1N1 influenza virus and control lung samples and to determine whether they could be clearly separated simply based on the type of infecting pathogen and to identify factors that might confound differential expression analysis. Principal components (PCs) capture orthogonal dimensions of variability in the data in descending order of contribution. Non-infected samples clearly separate from other samples on PCs 1 and 3, but this was not the case for SARS-CoV-2 and pH1N1 influenza samples (Fig. 2a). Indeed, across PCs 1-4 SARS-CoV-2 and pH1N1 influenza samples were substantially overlapping. These data suggest that the transcriptomic profile of SARS-CoV-2 and pH1N1 influenza virus tissues were not highly variable at low viral load, and that a common transcriptome was induced by these two viruses.

**Figure 2.**
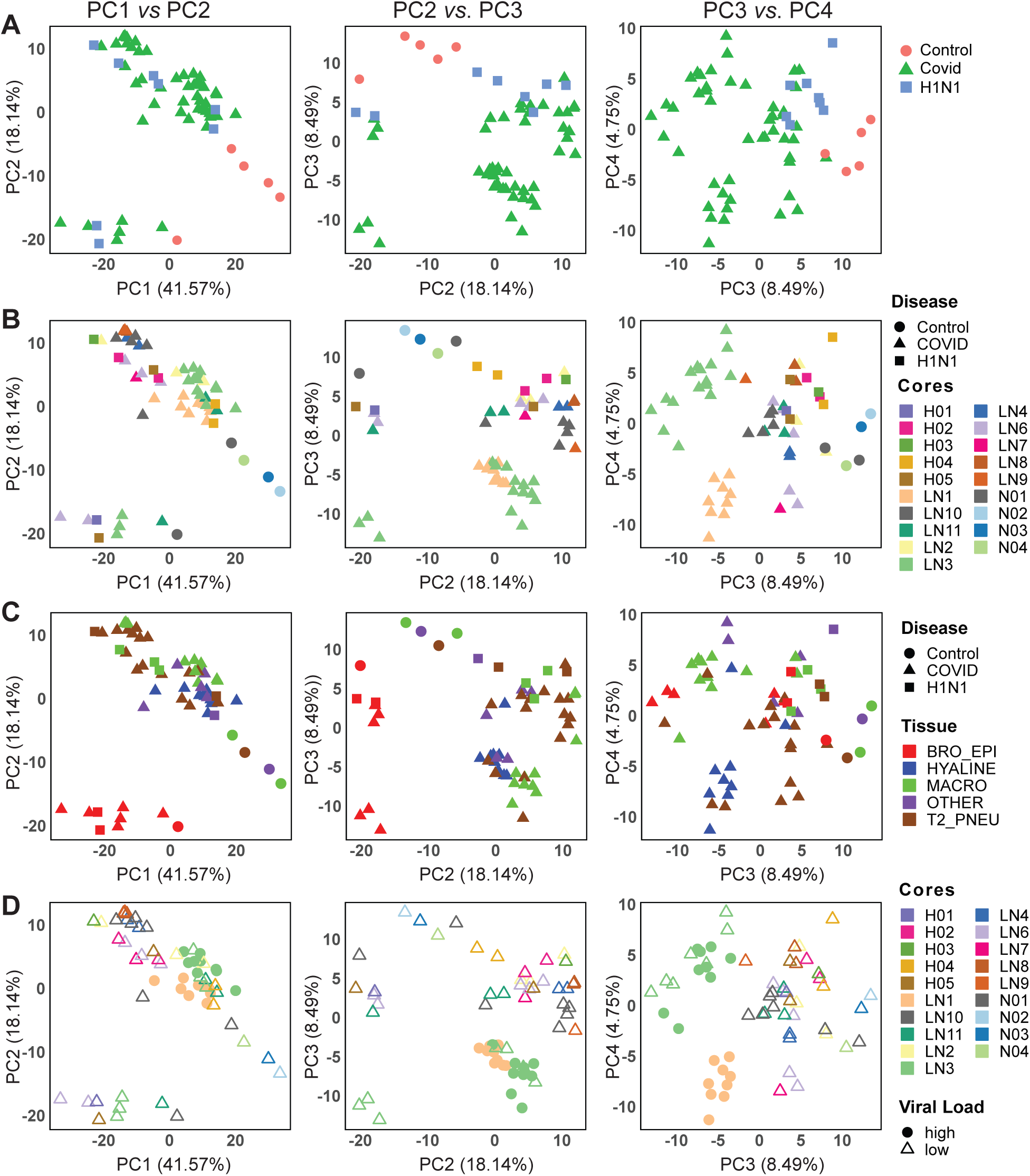
Principal components identify variability and factors in the transcriptomic data. Principal components (PCs) capturing orthogonal dimensions of variability in the transcriptomic data in descending order of contribution (i.e. PC1-PC2, PC3-PC2 and PC3-PC4) were plotted stratifying based on the following factors in the experimental data: (**a**) Disease types, (**b**) cores/patients with disease types, (**c**) dominant tissue types with disease types and (**d**) Viral Load in cores/patients. Disease type in panels **a, b** and **c** labelled as shapes with Control as circles, COVID as triangles and H1N1 as squares. Viral load labelled in panel **b** as shapes with high viral load as solid circle and low viral low as unfilled triangle. Cores and patients labelled in panel **b** and **c** using differing colors. Dominant tissue types in panel **c** labelled as differing fill colors (BRO_EPI, bronchiolar epithelium; HYALINE, hyaline membranes; MACRO, macrophages; T2_PNEU, Type2 pneumocytes; OTHERS, other classifications).

To further examine the major sources of variation in the dataset, we labelled PCA plots by multiple different parameters including sample cores (Fig. 2b), dominant cell type (Fig. 2c) and viral load (Fig. 2d). Plotting PC 3 and 4, and labelling by sample core revealed some patient effects - a clear separation of ROIs related to cores LN1 and LN3, and some clustering for other cores with multiple ROIs (LN10)(Fig. 2b). This suggests that within patient correlations and patient-patient variation has an important underlying effect on the observed transcriptome and that this needs to be taken into account in any subsequent analysis This analysis allowed us to determine that the samples clearly separating on PC2, and clearly clustered from other samples when plotting PC1 and 2, were grouped based on the dominant cell type found within the sample core (Fig. 2c). Specifically, cores where bronchiolar epithelial cells were the dominant cell type clustered together independent of whether the patient had pH1N1 influenza virus or SARS-CoV-2 (Fig. 2c), or belonged to the non-infected controls. This implied that the tissue heterogeneity captured by dominant cell type was a substantial contributor to the variability in our experiment. While epithelial cells are the primary cellular target of both SARS-CoV-2 and influenza virus^12^, our data did not identify viral epithelial tropism as a key regulator of signal as the cores dominated by bronchial epithelial cells did not exhibit the highest viral load (as defined by RNA Scope for SARS-CoV-2)(Fig. 2d). However, the high SARS-CoV-2 viral load cores separate from other virally-infected samples and control samples on PC3 (Fig. 2d). Together, these analyses suggest that unbiased and accurate comparative transcriptomics analyses between uninfected, SARS-CoV-2 and pH1N1 influenza lung samples must take account of the dominant cell type present in samples, inter-patient variations and within-patient correlations. The ability to include structural and spatial covariates demonstrates the key advantage in using a spatial transcriptomics as compared with bulk RNA sequencing, but also necessitates careful construction of the model used to compare expression across the ROIs sampled.

### Severe COVID-19 infection results in a global pro-inflammatory response and downregulation of immune cell effector and regeneration pathways

Differential gene expression analysis was subsequently performed on all sample cores, taking into account the effect of the dominant cell type and the appropriate within and inter-patient variations. 286 genes were identified as significantly upregulated and 20 genes were significantly downregulated in the lungs of SARS-CoV-2 patients compared with non-virally infected controls (Fig. 3, Extended Data Table 6, 7).

**Figure 2.**
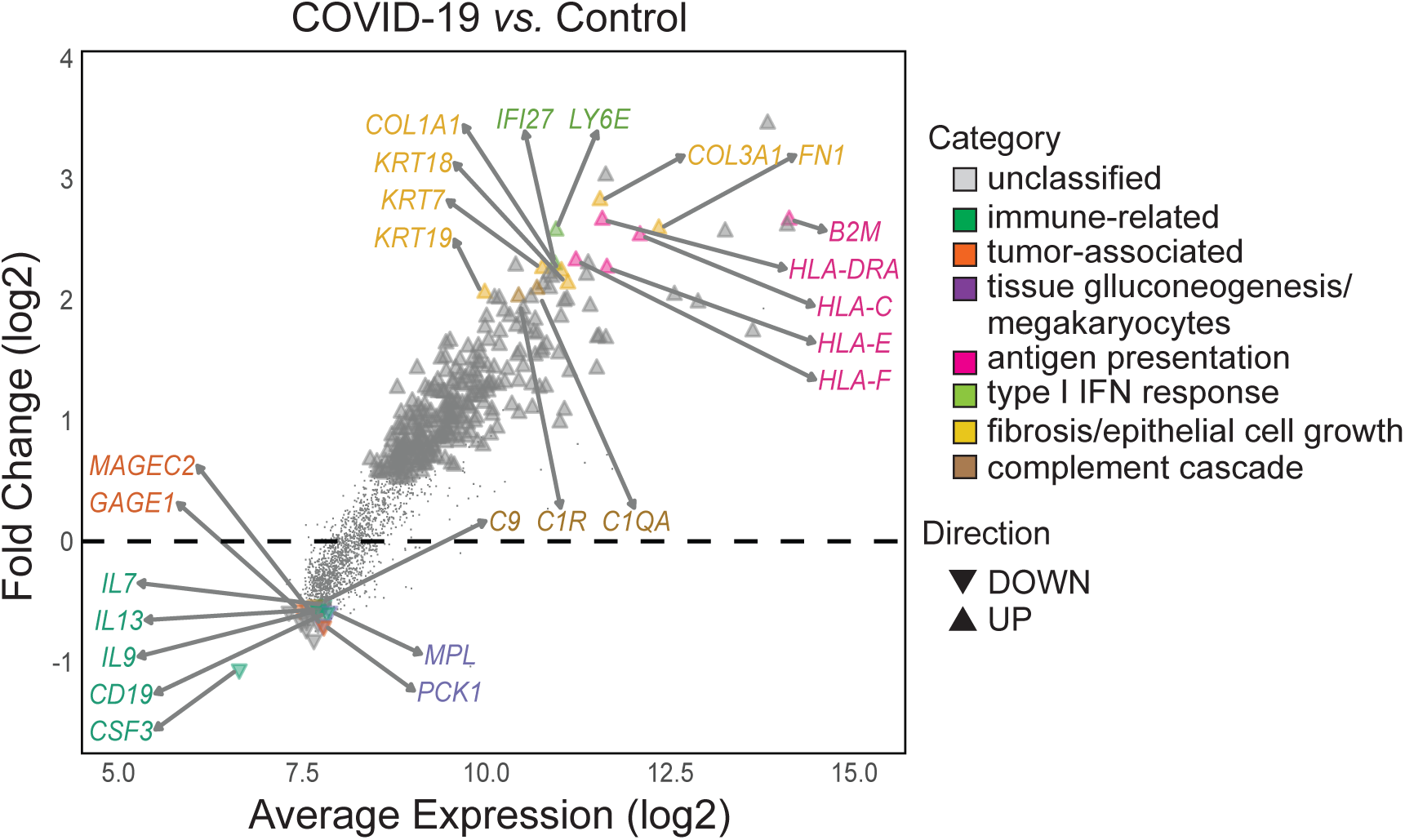
COVID-19 infection drives pro-inflammatory response and suppresses immune cell effector and regeneration. Distribution of differentially expressed genes as a function of the average transcript expression and fold change (log2) identified in COVID-19 samples *vs* uninfected (control) samples. Down-regulated genes included immune related, cytokine, tumor/cell-survival associated genes and cell regeneration genes, inferring a suppression of immune cell effect and regeneration. Up-regulated genes are enriched with pro-inflammatory genes including Type I interferon (IFN) response and fibrosis genes. Representative genes from these processes are highlighted. Differential expression genes generated using the *voom-limma* pipeline with *limma:duplicationCorrelations* and applying TREAT criteria with absolute log fold change > 1.2 with p-value <0.05.

**Figure 3.**
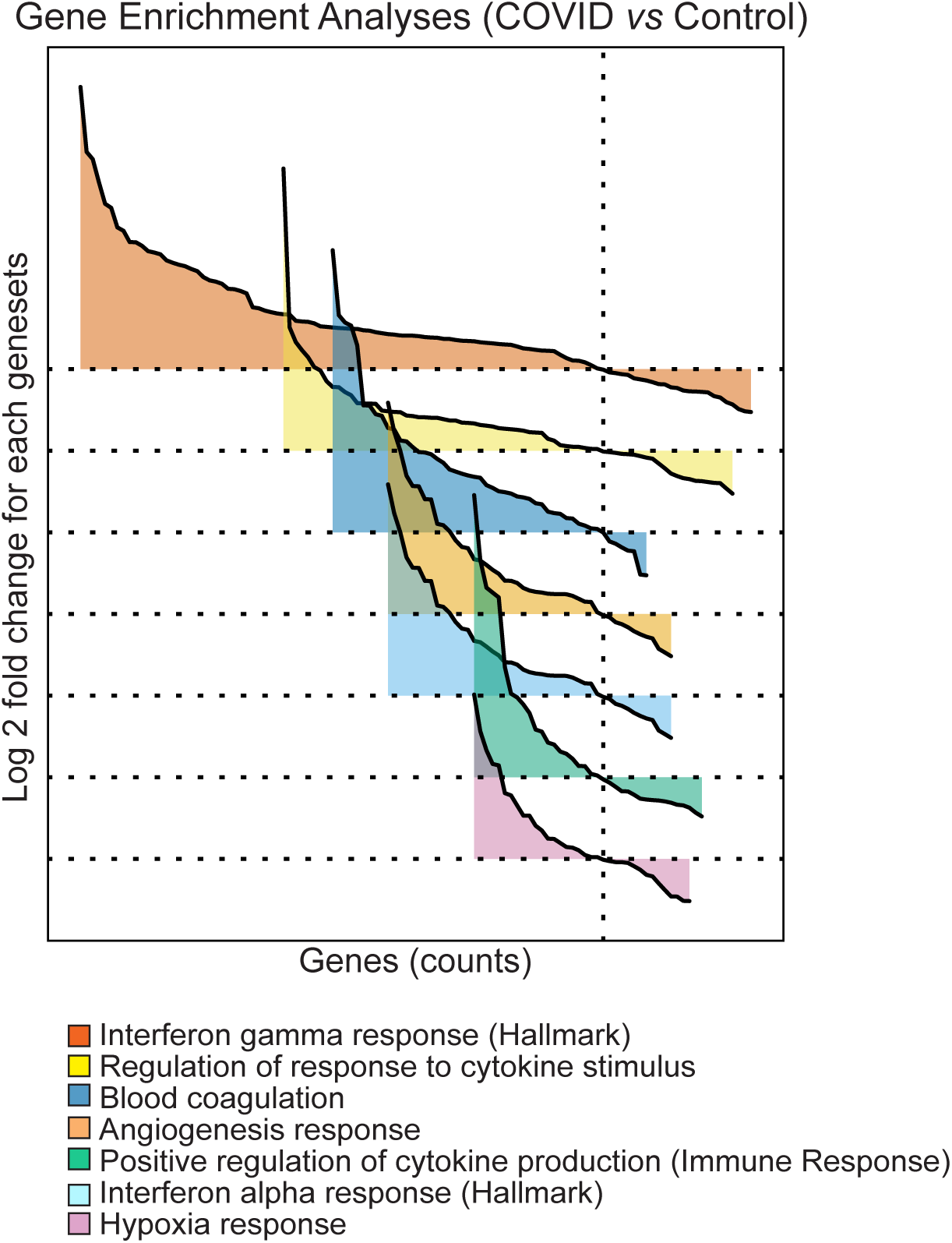
Gene set enrichment analyses (GSEA) reveals upregulated cytokine responses with accompanying coagulopathies in COVID-19 infection. GSEA for differentially expressed genes comparing COVID-19 and uninfected samples were conducted using the estimated log2 fold change and p-values with the distribution of significantly differentiated genesets visualised as a function of the log2 fold change (logFC) and genes sorted by the logFC. The genesets are custom genesets for (**a**) angiogenesis response, (**b**) blood coagulation (**g**) hypoxia responses based on nanoString’s nCounter® PanCancer Progression Panel were identified by GSEA to be differentially upregulated in COVID-19 samples. (see *Table* 2). (**e, f**) MSigDB Hallmark gene set for IFN-α and IFN-γ. (**c, d**) MSigDB Gene Ontology (GO), biological processes (BP) collection identified by GSEA for cytokine regulation and responses. Refer to comprehensive list in Table 1. GSEA conducted using limma-fry with FDR <0.05. The direction and relative change in expression are shown. Details of each geneset are provided in Extended Data Figure 4.

Down regulated genes included immune-related genes such as B lymphocyte antigen *CD19*, cytokine genes such as *IL7, IL9* and *IL13* and components of the complement cascade (e.g. C9) together with *CSF3*, colony stimulating factor, normally associated with mobilization of cells from the bone marrow (Extended Data Table 6). A number of tumor-associated genes including *GAGE1* and *MAGEC2* were downregulated, with these genes being associated with cell survival. Interestingly, *PCK1*, which is required for tissue gluconeogenesis, and *MPL*, that regulates the generation of megakaryocytes and thus platelet formation were also highly reduced^13^.

A more diverse array of upregulated genes was found to be differentially expressed in the lungs of SARS-CoV-2 patients compared with those from patients who died of non-viral causes (Extended Data Table 7). These included genes associated with antigen presentation (e.g. *B2M, HLA-DRA, HLA-C* and *HLA-E)*, the Type I interferon (IFN) response (e.g. *LY6E* and *IFI27*), fibrosis and epithelial cell growth (e.g. *COL3A1, FN1, KRT7, COL1A1, KRT18 and KRT19)* and the complement cascade (*C1QA* and *C1R*). Consistent with these observations, gene set enrichment analysis showed that hallmark genes of biological processes such as reactive oxygen species, complement, IL6/JAK/STAT3 signalling, apoptosis. p53 signalling IFN-γ response, hypoxia, IFN-α response, coagulation, TGF-β signalling, IL-2/STAT5 signalling, angiogenesis, TNF-α signalling via NFκ-B, and inflammatory response were all significantly upregulated in SARS-CoV-2 infected lungs (Table 1). Gene set enrichment using blood coagulation, hypoxia responses and angiogenesis related genes from nanoString’s nCounter® PanCancer Progression Panel further confirmed the upregulation of pathways associated with blood coagulation and angiogenesis responses (Table 2). These analyses show that SARS-CoV-2 samples significantly upregulation genes associated with blood coagulation, angiogenesis, the IFN-α and IFN-γ response and positively regulate cytokine production and cytokine stimulus during the immune response (Fig. 4). These data are consistent with previous descriptions of a ‘cytokine storm’ response in patients with severe SARS-CoV-2, often accompanied by various coagulopathies^14^. They are also in agreement with the notion that a late stage or delayed type I IFN response may be observed in concert with a pro-inflammatory response^15^.

**Figure 4.**
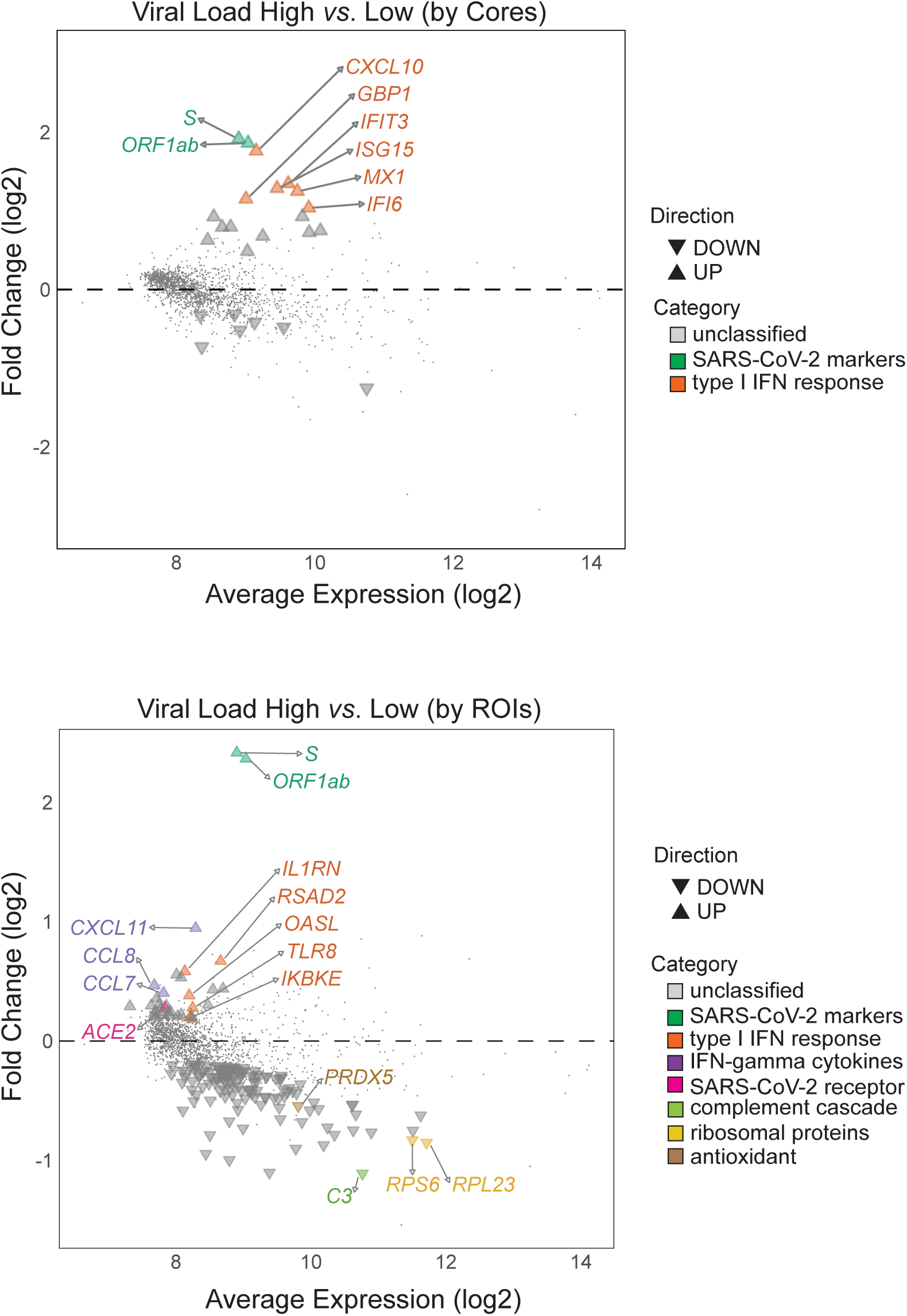
Resolution provided by spatial data reveals type I interferon gene signature associates with COVID-19 viral load. Distribution of differentially expressed genes as a function of the average transcript expression and fold change (log2) identified in high viral load vs low viral load COVID-19 samples. Analyses were conducted at two resolutions, firstly by (**a**) grouping ROIs from the same cores with the same degree of viral load (core patients-based approach); secondly by (**b**) treating each regions/ROIs sample as an independent observation (ROI-based approach). In both approaches, SAR-CoV-2 specific genes *S* and *ORF1ab* were strongly upregulated in the high viral load group, consistent with results of the RNAscope. Consistent with previous reports, type I IFN response genes were associated with SAR-CoV-2 RNA expression. Notably the core-based approach reveals limited differential gene expression with the finer resolution ROI-based approach revealing additional differential changes in complement cascade, ribosomal protein and antioxidants genes. Representative genes are highlighted. Differential expression genes derived using *voom-limma* pipeline with *limma:duplicationCorrelations* and applying absolute log2 fold change > 1.0 with p-value <0.05.

### Limited differential gene expression associated with SARS-CoV-2 viral load

The pathogenesis of SARS-CoV-2 is a complex interplay of host immunopathology and viral load. To determine how viral load influenced the host transcriptomic signal, we performed a differential expression analysis of SARS-CoV-2 samples stratifying based on viral load. Patient sample cores were classified as either ‘high’ or ‘low’ based on the presence of viral mRNA by RNAscope^®^. Thus, two cores which were scored positively by RNAscope^®^, LN1 and LN3, were designated as ‘high’ while the remaining 8 cores from SARS-CoV-2 patients were scored as ‘low’. This analysis showed that only 17 up-regulated and 7 down-regulated genes were differentially expressed between the high viral load SARS-CoV-2 infected cores and the other SARS-CoV-2 cores (Fig. 5, Table 3). Consistent with our RNAScope analyses, SAR-CoV-2 specific genes *S* and *ORF1ab* were the most strongly upregulated genes in the high viral load group (1.916 and 1.861 log2 fold change, respectively). All other upregulated genes with >1 log2 fold change in expression were associated with the type I IFN response (*CXCL10, IFIT3, ISG15, MX1, GBP1* and *IFI6*)(Fig. 5a, Table 3).

**Figure 5.**
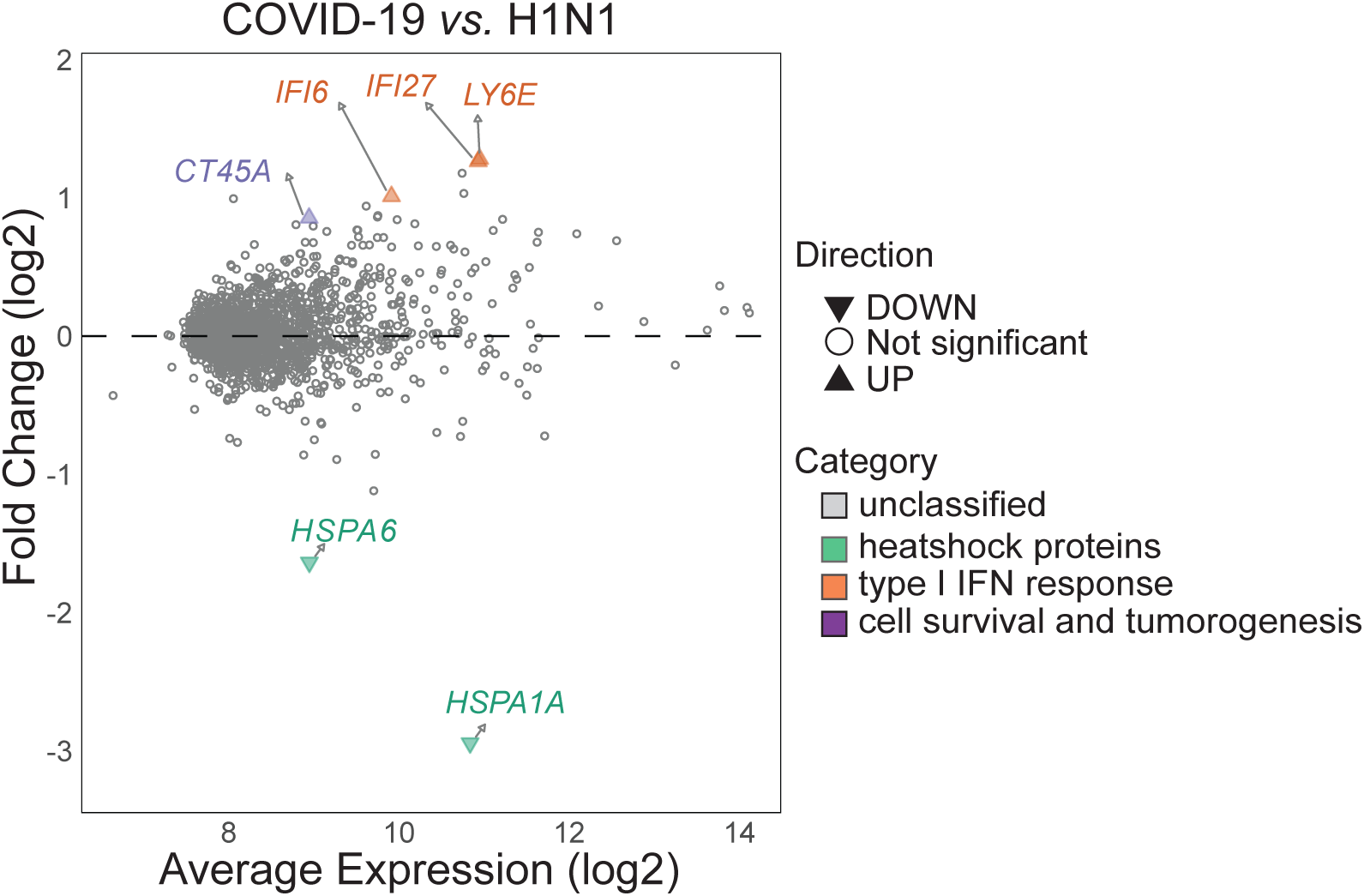
Limited transcriptomic differences associated with COVID-19 infection compared with H1N1. Distribution of differentially expressed genes as a function of the average transcript expression and fold change (log2) identified in COVID-19 samples vs pH1N1 samples were visualised with the 6 differentially expressed genes labelled and classified based on associated biological processes. The genes associated with the type I Interferon response, heat shock protein family members and (associated) with cell survival and tumorigenesis are shown. This exclusive list of genes reveals the subtle differences between COVID-19 and H1N1 infected transcriptome and presents a potential disease specific transcriptomic signature for distinguishing the two disease types. Differential expression genes derived using *voom-limma* pipeline with *limma:duplicationCorrelations* and applying *TREAT* criteria with absolute log fold change > 1.2 with p-value <0.05.

The above analysis assumed that every ROI derived from the same core would have the same degree of viral infection (i.e. high or low). However, viral infection is unlikely to be uniform across tissues. Indeed, none of the bronchiolar epithelial ROIs in the two viral high cores were classified as high (Fig. 2d, Extended Data Table 5). To examine the impact of this, we considered each ROI as an independent sample which was classified as either ‘high’ (score>0) or ‘low’ by RNAscope positivity. By stratifying cores with this criterion and accounting for the dominant cell type as well as correlations between ROIs from the same core, we identified 33 differentially upregulated genes and 132 downregulated genes in high viral load ROIs compared to low viral load ROIs (Extended Data Table 8). Once again, *S* and *ORF1ab* were the most strongly upregulated genes in the high viral load group (2.415 and 2.365 log2 fold change respectively). Other upregulated genes of note were those associated with the type I IFN response (e.g. *RSAD2, OASL, TLR8, IL1RN* and *IKBKE*), chemokines associated with IFN−γ (e.g. *CXCL11, CCL8* and *CCL7*) and the SARS-CoV-2 receptor *ACE2*. A more diverse array of genes were downregulated in high viral load samples including those associated with the complement cascade (e.g. *C3*), ribosomal proteins (e.g. *RPS6* and *RPL23*) and antioxidants (e.g. *PRDX5*)(Figure 5B). Taken together, these data suggest that a pronounced type I interferon gene signature is associated with SARS-CoV-2 RNA expression and the spatial context provided by this technology enables finer details of viral load associations to be determined.

### Severe COVID-19 is associated with a limited, differential transcriptome when compared with severe influenza

Only a limited number of genes were identified as differentially expressed between COVID-19 and pH1N1 influenza samples (2 downregulated and 4 upregulated genes, Fig. 6). Of the six genes significantly differentially regulated in COVID-19 samples, three were associated with the type I IFN response (*LY6E, IFI27* & *IFI6*), two were heat shock protein family members (*HSPA6* & *HSPA1A*) and one, *CT45A1*, when combined with various growth factors, is associated with cell survival and tumorigenesis. Interestingly, all three of the interferon-inducible genes were also significantly upregulated in COVID-19 samples compared to those without a viral infection (Extended Data Table 7). Two heat shock protein family members (*HSPA1A* and *HSPA*) were also identified as significantly down-regulated in COVID-19 tissues. These two proteins were found to be significantly upregulated in pH1N1 tissues compared to uninfected tissues (data not shown), suggesting a potential influenza specific signature. Together, these data indicate that pH1N1 influenza virus and SARS-CoV-2 induce only subtly different host pulmonary transcriptomes but may be potentially distinguished by a core disease-associated immune signature.

## Discussion

Understanding the biological functions, networks, and host-pathogen interactions that impact organ and disease development requires both cellular information and a spatial context. Analyses of bulk RNA sequencing or scRNA sequencing provides a global overview of an organ’s response to a pathogen, typically identifying broad inflammatory pathways. However, such approaches fail to identify subtle individual cellular changes that are spatially distinct. This has particular impact when considering innate and adaptive immune cells that may be crucial for understanding pathogen-specific responses, and to distinguish the profile of one pathogen from another. In contrast, spatially resolved transcriptomes of virally-infected tissues offers the possibility of disentangling the individual infected cells, contributions of viral load, cellular responses and hence patient-to-patient variability.

The clinical spectrum of COVID-19 is highly variable. Association of viral load with disease severity would provide a mechanism for early stratification of patients for treatment options. Indeed, analyses of nasopharyngeal swabs suggests that nasopharyngeal SARS-CoV-2 RNA is independently correlated with disease severity^16^, similar to earlier observations with SARS-CoV-1^17^. However, the association between viral replication in the lung and severe disease remains more complicated. Here, we observed that 8 out of 10 patients who died because of COVID-19 had no detectable viral RNA in lung biopsies as determined by RNAscope. Transcriptomic analysis showed that areas of high viral load were associated with a pronounced type I IFN response, consistent with other preliminary spatial analysis of the lungs of COVID-19 patients ^11^ and assumedly due to increased viral pathogen-associated molecular patterns (PAMPs) available for type I IFN stimulation. Nevertheless, even in samples where no viral RNA was detected, a pronounced type I IFN gene signature could still be observed in the pulmonary tissue. These observations add weight to the growing understanding of the role type I IFNs in SARS-CoV-2 infections^18^. Current evidence suggests that an early and robust induction of type I interferons can help control viral replication and help ensure a mild infection. In contrast, a delayed induction of type I IFN (i.e. after the peak of viral replication) is largely irrelevant for viral control as viral titres have already declined at this point in the infection but may help perpetuate the detrimental pro-inflammatory response and lung damage ^18,19^. The transcriptomic data presented here thus speak to the nuanced role of type I IFNs in SARS-CoV-2 infection.

Pulmonary transcriptomic analysis is a powerful tool to delineate the pathogenesis of SARS-CoV-2 and identify how it differs compared to other respiratory pathogens such as influenza virus. Previous studies have suggested that the lungs of COVID-19 patients display an increased incidence of alveolar capillary microthrombi and thrombosis with microangiopathy compared to those of influenza patients^5^. In the present study, D-dimer levels were elevated in six of ten COVID-19 patients and some thrombosis was observed. Consistent with these observations, an earlier report using bulk lung RNA analyses to identify 69 differentially-expressed angiogenesis-related genes in COVID-19 patients, but not in influenza patients^11^. Strikingly, while we observed that both COVID-19 and influenza patients had an upregulation of genes associated with coagulation and angiogenesis, these genes were not differentially expressed between the two virus-infected patient groups. These contrasting data point to the complementary role of blood profiling, bulk and tissue-specific spatial analyses in defining clinical parameters for therapy.

Accurate detection, timely diagnosis and effective treatment are all essential to the management of COVID-19 and will be instrumental in curbing viral spread as the number of new infections increase daily. Several potential clinical and transcriptional biomarkers for triaging patients with COVID-19 have been explored. Clinical biomarkers include C-reactive protein, serum amyloid A, interleukin-6, lactate dehydrogenase, neutrophil-to-lymphocyte ratio, D-dimer, lymphocytes and platelet count ^20,21^. However, the majority of these studies have focussed on biomarkers in patient blood. While peripheral blood samples are a convenient and highly accessible site for clinical sampling, this is not the primary site of viral infection. Thus, information delineated from analyses of blood may not reflect tissue severity and thus may hamper biomarker discovery. Furthermore, a biomarker that could be rapidly identified in the respiratory tissue (e.g. *via* a nasal swab) may be more accessible in low resource settings than one requiring a blood sample. Interestingly, previous analyses of *IFI27* (interferon-alpha inducible protein 27) in blood of COVID-19 patients revealed that *IFI27* was upregulated in SARS-CoV-2 infection^22-24^. Here, we identified *IFI27* as differentiated upregulated in the lungs of both COVID-19 patients *vs* control patients and in the highly restricted set of genes differentially expressed between COVID-19 and influenza patients. *IFI27* has previously been used as a biomarker to successfully differentiate bacterial pneumonia from influenza virus infection^25^. These data raise the exciting possibility that *IFI27* may not only represent a biomarker for severe COVID-19 but that it may also help differentiate this disease from other clinically similar viral infections. This becomes particularly important as the ‘second wave’ of COVID-19 in the US and Europe is set to overlap with the winter influenza season. Validation of this gene in nasal specimens, as well associations with disease severity will be required to confirm *IFI27* as a gene signature that is useful in stratifying COVID-19 patients.

This study was subject to several important limitations. Firstly, all data was derived from a small sample cohort derive from a single study site, and it remains to be determined how much these data can be extrapolated to other patient populations. Furthermore, additional studies are required across a broader range of patients (i.e. those with mild and moderate disease) to determine the therapeutic value of any of the putative tissue biomarkers identified herein. However, despite these limitations these data reveal the unprecedented power of spatial profiling combined with detailed multiparameter bioinformatic analyses to dissect the key variables that contribute to differential gene expression across highly variable patient cohorts and the heterogeneous distribution of virus and immune responsiveness within tissues. This study also demonstrated the value of using the suite of linear-modelling tools available in *limma* to interrogate the complex multi-factorial experiment design in this study. In particular, *limma*’s ability to model complex experimental designs and to implement information borrowing amongst genes to handle the relatively small number of samples in the panel, make it a highly attractive analysis tool for spatial profiling techniques with targeted gene panels.

The present study suggests that spatial profiling would present many advantages in analysing COVID-19 samples across different patient cohorts to identify fundamental response signatures distinct from background effects. This work therefore established the foundation to evaluate larger tissue cohorts to assess the relationship between blood and tissue biomarkers, disease progression, pathology and repair.

### Data and materials availability

Nanostring data that support the findings of this study will be deposited with the Gene Expression Omnibus (GEO) repository and made available on publication. All the data to evaluate the conclusions in this paper are present either in the main text or in supplementary materials.

## Data Availability

Nanostring data that support the findings of this study will be deposited with the Gene Expression Omnibus (GEO) repository and made available onpublication. All the data to eval uate the conclusions in this paper are present either in the main text or in supplementary materials.

## Acknowledgments

We thank S. Weaver, M. Haraguchi (Fred Hutch Experimental Histopathology Department, USA), L. Pan, A. Nam (Nanostring Technologies Seattle, USA), I. Stefani (University of Queensland Diamantina Institute Translational Innate Immunology Group) and G. Smyth (WEHI). We thank the patients and their families that made this study possible.

## Author Contributions

AK, FSFG, KRS, GTB, KOB conceived of the study. AK, AFRDSM, JM, JDSMJ, CBVDP,

SN, CPB, PSFG, LDN, CC, TM, GRR performed the experimentation. AK, CWT, DB, JM, CC, BT, KRS, MJD, FSFG, GTB performed the data analysis and interpretation. All authors critically reviewed and approved the manuscript prior to submission.

## Methods

### Patient selection

Tissue microarray (TMA) cores were prepared from autopsied pulmonary tissue from 10 SARS-CoV-2 and 5 pH1N1 patients who died from respiratory failure (ARDS)(Extended Data Table 1-3). Control material was obtained from 4 uninfected patients (Extended Data Table 3). All SARS-CoV-2 and pH1N1 patients were confirmed for infection through RTqPCR of nasopharyngeal swab specimens, and imaging with computed tomography (CT) showed diffuse and bilateral opacities with ground-glass attenuation, suggestive of viral pulmonary infection. Autopsy and biopsy materials were obtained from the Pontificia Universidade Catolica do Parana PUCPR the National Commission for Research Ethics (CONEP) under ethics approval numbers 2020001792/30188020.7.1001.0020 and 2020001934/30822820.8.000.0020. The study was also approved under University of Queensland Human Research Ethics Committee (HREC) ratification.

### Tissue Preparation and Histopathology

Thirty 5µm thick serial sections were cut from the TMA blocks onto positively charged slides (Bond Apex) and sections were stained with hematoxylin and eosin (H&E) and Masson’s trichrome stain. Brightfield images were obtained using an Aperio (Leica Biosystems, US) slide scanner for assessment of histopathology by a pathologist. TMA cores were assessed for overall histological pattern. Regions of interest (ROIs) were semi-quantitatively analysed for alveolar haemorrhage and oedema, hyaline membrane formation, acute inflammation, type 2 pneumocyte hyperplasia, squamous metaplasia, capillary congestion, fibroblastic foci, and interstitial inflammation.

### Immunohistochemistry and RNAscope®

Immunohistochemistry was performed on a Leica Bond-RX autostainer (Leica Biosystems, US) with antibody targeting SARS-CoV-2 spike protein (Abcam, ab272504) at 2 μg/ml. Heat induced epitope retrieval was performed in buffer ER1 at 100°C for 20 minutes, and signal visualized with 3,3’-Diaminobenzidine (DAB) substrate. Slides were imaged using a Zeiss Axioscanner (Carl Zeiss, Germany) and IHC was scored by a pathologist for bronchiolar epithelium, type 2 pneumocytes, interstitial lymphocytes and alveolar macrophage compartments.

RNAscope^®^ probes (ACDbio, US) targeting SARS-CoV-2 spike mRNA (nCoV2019, #848561-C3), ACE2 host receptor mRNA (#848151-C2), and host serine protease TMPRSS2 mRNA (#470341-C1) were used as per manufacturer instructions for automation on Leica Bond RX. DNA was visualised with Syto13 (Thermofisher Scientific), channel 1 with Opal 570 (1:500), channel 2 with Opal 620 (1:1500), and channel 3 with Opal 690 (1:1500)(PerkinElmer). Fluorescent images were acquired with Nanostring Mars prototype DSP at 20x.

### Nanostring GeoMX DSP Covid-19 Immune Atlas Panel

Freshly cut sections of each TMA were processed according to the Nanostring GeoMX Digital Spatial Profiler (DSP) slide preparation manual by the technology access program. Briefly, slides were baked 1 hour at 60°C and then processed by Leica Bond RX autostainer. Slides were pre-treated with Proteinase K and then hybridised with mRNA probes contained in the Covid-19 Immune Atlas panel with additional SARS-CoV-2 spike-in panel. After incubation, slides were washed and then stained with CD68, CD3, PanCK, and Syto83 for 1 hour then loaded into GeoMX DSP instrument for scanning and ROI selection. Selected ROIs were guided by RNAscope^®^ and IHC positivity on SARS-CoV-2 tissues with similar tissue structures captured on H1N1 and uninfected tissue cores. Oligonucleotides linked to hybridised mRNA targets were cleaved and collected for counting using Illumina i5 and i7 dual indexing. PCR reactions were performed with 4 μl of a GeoMx DSP sample. AMPure XP beads (Beckman Coulter) were used at 1.2× bead-to-sample ratio for PCR product purification. Paired-end sequencing (2×75) was performed using NextSeq550 up to 400M total aligned reads. Fastq files were processed by DND system and uploaded to GeoMX DSP system where raw and Q3 normalized counts of all targets were aligned with regions of interest (ROIs).

### Transcriptomic Data

Data used in this study result from an mRNA assay conducted with the NanoString’s GeoMx Covid-19 Immune atlas panel using the GeoMX Digital Spatial Profiler (DSP). The data were measurements of RNA abundance of over 1800 genes, including 22 add-in COVID related genes, 4 SARS-CoV-2 specific genes and 2 negative control (SARS-CoV-2 Neg, NegProbe) genes and 32 internal reference genes. Samples were acquired from 3 TMA slides each containing 10 tissue cores from 10 patients. The three slides correspond to either COVID-19, H1N1 or Control Tissue biopsies. Transcriptomic measurements were made on regions of interests within each core. Control samples are COVID-free and H1N1-free, but they are not necessarily disease-free samples; they originate from patients with other non-viral diseases. In total, 60 ROIs were analysed (47 COVID-19, 8 H1N1 and 5 “Control”). Factors considered in this dataset include disease type (COVID, H1N1 and Control), patient of origin, dominant tissue type (Type 2 pneumocytes, bronchiolar epithelium, hyaline membranes, macrophages) and viral load (based on RNAscope, RNAscope scores).

### Bioinformatic analyses

Data exploration and quality checks were conducted on the Q3 normalised count data generated from the CTA DSP mRNA assay. Relative log expression (RLE) plots were used assess the presence of unwanted variation in the form of batch effects ^26^. Data were normalised using the trimmed mean of M-values (TMM) method ^27^ using the all the genes in the panel. Specifically, log-transformed transcript abundance data were median-centred for each gene, and then within each sample the difference between the observed and population median of each gene was calculated. Principal components analysis (PCA) of the samples was conducted to identify variability related to specific factors in the dataset and experimental design.

Differential expression (DE) analysis, Gene Ontology and KEGG pathway enrichment analysis were carried out using R/Bioconductor packages edgeR (v3.30.3)^28,29^ and limma (v3.44.3)^30^. Briefly, differential expression was modelled using linear models with various experimental factors as predictors. Variation in gene expression was modelled as the combination of a common dispersion that applies to all genes and a gene-specific dispersion. Limma was used to model and estimate the variation of each gene by borrowing information from all other genes using an empirical Bayes approach, thereby allowing estimation of the common and gene-wise variation with as few as 2 replicates per condition. Once the linear model was fit to a given experimental design, various contrasts of interest were used to query the data for differential expression. The resulting statistic was an empirical Bayes moderated t-statistic which was more robust than a t-statistic from a classic t-test. Based on the observed differences between cores and tissue structures from the PCA analyses, considerations were made to allow for similarity that exists for regions originating from the same core using *duplicationCorrelations* in limma^31^. The flexible modelling framework afforded by linear models was used to take into account differences between heterogenous tissue structures (i.e. bronchiolar epithelial samples *vs* the rest of the samples) by including them as covariates in the models.

Two main factors were investigated in this analysis, namely disease type (COVID19, H1N1 and Control) and viral load. For disease type, two comparisons were investigated: COVID against *Control* with *cores* and *dominant tissue type* as covariates, and COVID against H1N1 samples, with *cores* and *dominant tissue type* as covariates. For these two contrasts, the voom-limma with *duplicationCorrelations* pipeline^32^ was used to fit linear models. The TREAT criteria was then applied^33^ (p-value <0.05) to perform statistical tests and subsequently calculate the t-statistics, log-fold change (logFC), and adjusted *p*-values for all genes. For viral load, the voom-limma with *duplicationCorrelations* pipeline^32^ (p-value <0.05) was used to fit linear models for the comparison between high and low viral load regions originating from COVID-19 patient cores (disease type as a covariate). The contrast was applied on two resolutions, firstly by grouping ROIs from the same cores with the same degree of viral load (cores/patients-based approach); secondly by treating each regions/ROIs sample as an independent observation (ROI-based approach). Gene set enrichment analysis (GSEA) were performed using the *fry* approach from the limma package. Gene sets from the Molecular Signatures Database (MsigDB) Hallmarks ^34,35^, C2 (curated gene sets) and C7 (immunologic signature gene sets) categories were tested using *fry*. Pathways from the KEGG pathway database were tested using the *kegga* function in the limma package and gene ontology enrichment was assessed using *goana* from the limma package^31^.

## Figure Legends

**Extended Data Figure 1.**
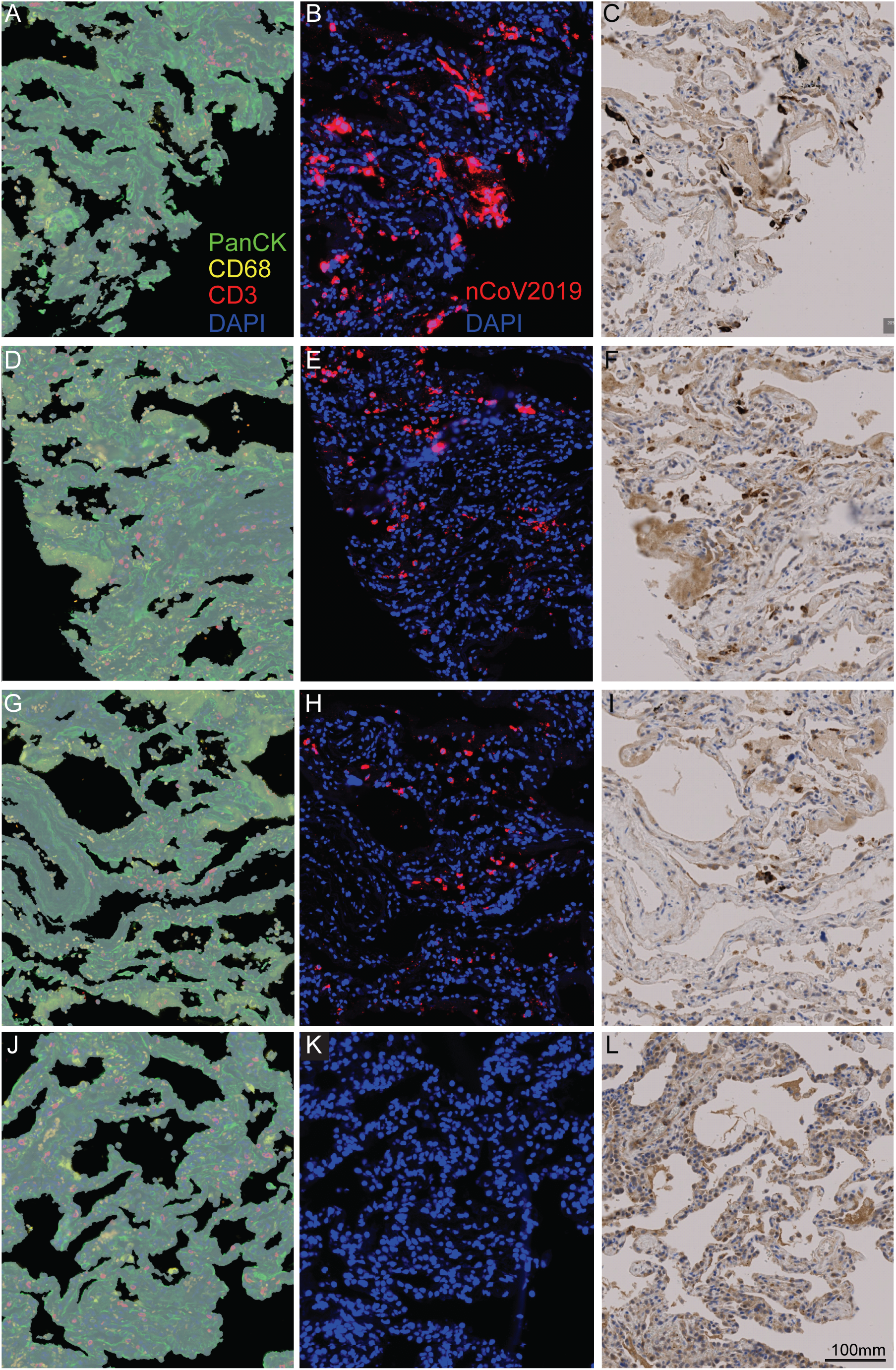
Representative assessment of virus load in DSP ROIs using RNAscope and IHC for SARS-CoV-2 spike mRNA and protein, respectively, on serial sections. Fluorescent IHC staining was performed for DSP acquisition using monocytes (CD68, yellow), T cells (CD3ε, red), lung epithelial cells (PanCK, green) and nuclei (DAPI, blue), (left panels, **A, D, G, J**). RNAscope staining was performed as per manufacturer’s instructions using nCoV2019-s probe for SARS-CoV-2 spike mRNA (center panels, **B, E, H, K**). IHC was performed with antibody against SARS-CoV-2 spike protein (right panels, **C, F, I, L**). ROIs were semi-quantitatively scored from 0 to 3 based on RNAscope signal abundance. **A-C**. Representative ROI with RNAscope score of 3 (high). **D-F**. Representative ROI with RNAscope score of 2 (intermediate). **G-I**. Representative ROI with RNAscope score of 1 (low). **J-L**. Representative ROI with RNAscope score of 0 (below detection). Scale bar = 100µm.

**Extended Data Figure 2.**
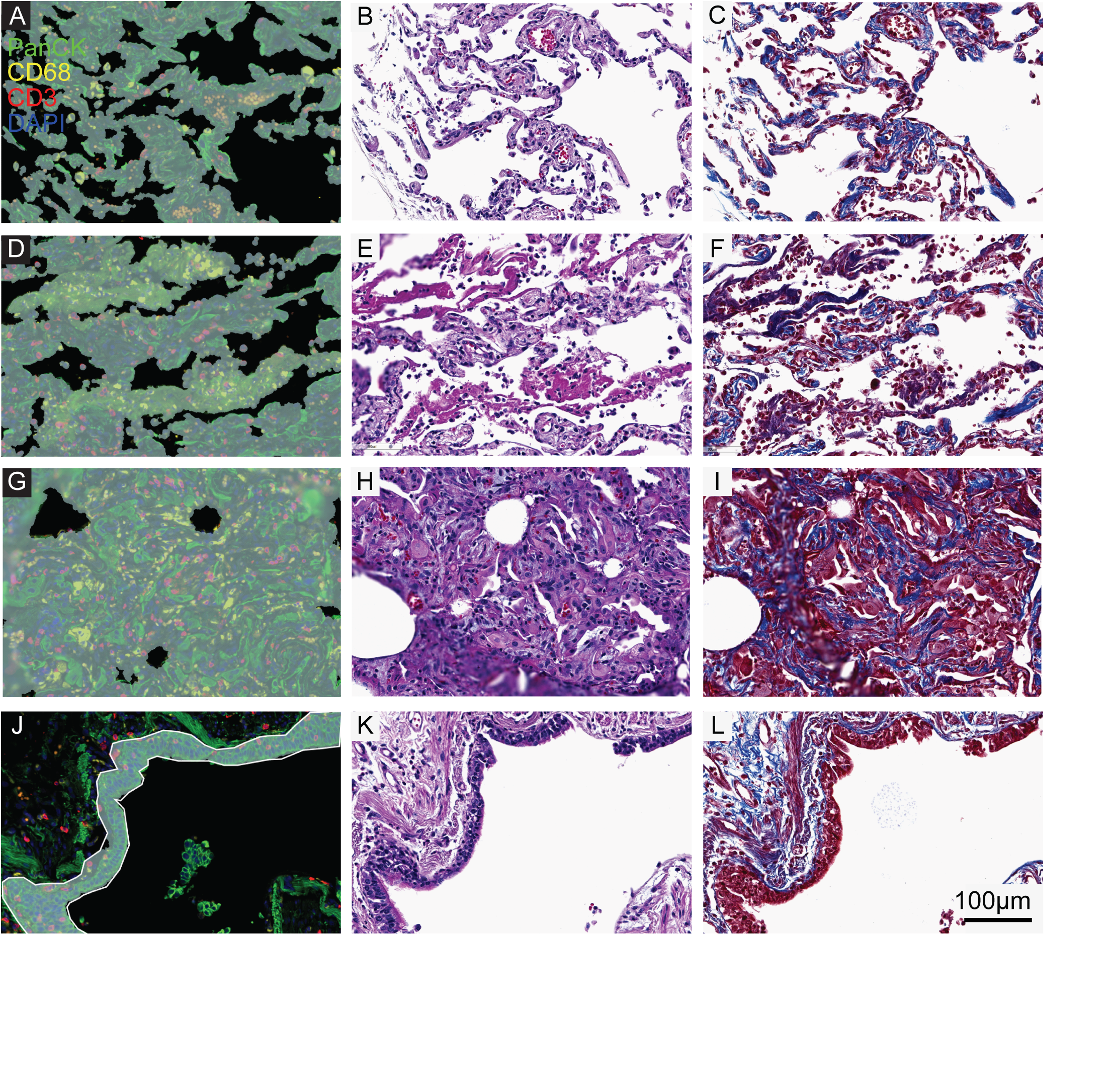
Representative assessment of dominant tissue histopathology in DSP ROIs. TMAs were stained for DSP acquisition using monocytes (CD68, yellow), T cells (CD3ε, red), lung epithelial cells (PanCK, green) and nuclei (DAPI, blue)(left panels). H&E (middle panels) and Masson’s trichrome (right panels) histological staining was performed on serial sections and ROIs overlaid for assessment by a pathologist. **A-C**. ROIs characterised by alveolar type 2 pneumocytes. **D-F**. ROIs characterised by hyaline membranes. **G-I**. ROIs characterised by macrophage infiltration. **J-L**. ROIs specifically drawn around bronchiolar epithelium lining greater airways. Scale bar = 100µm.

**Extended Data Figure 3.**
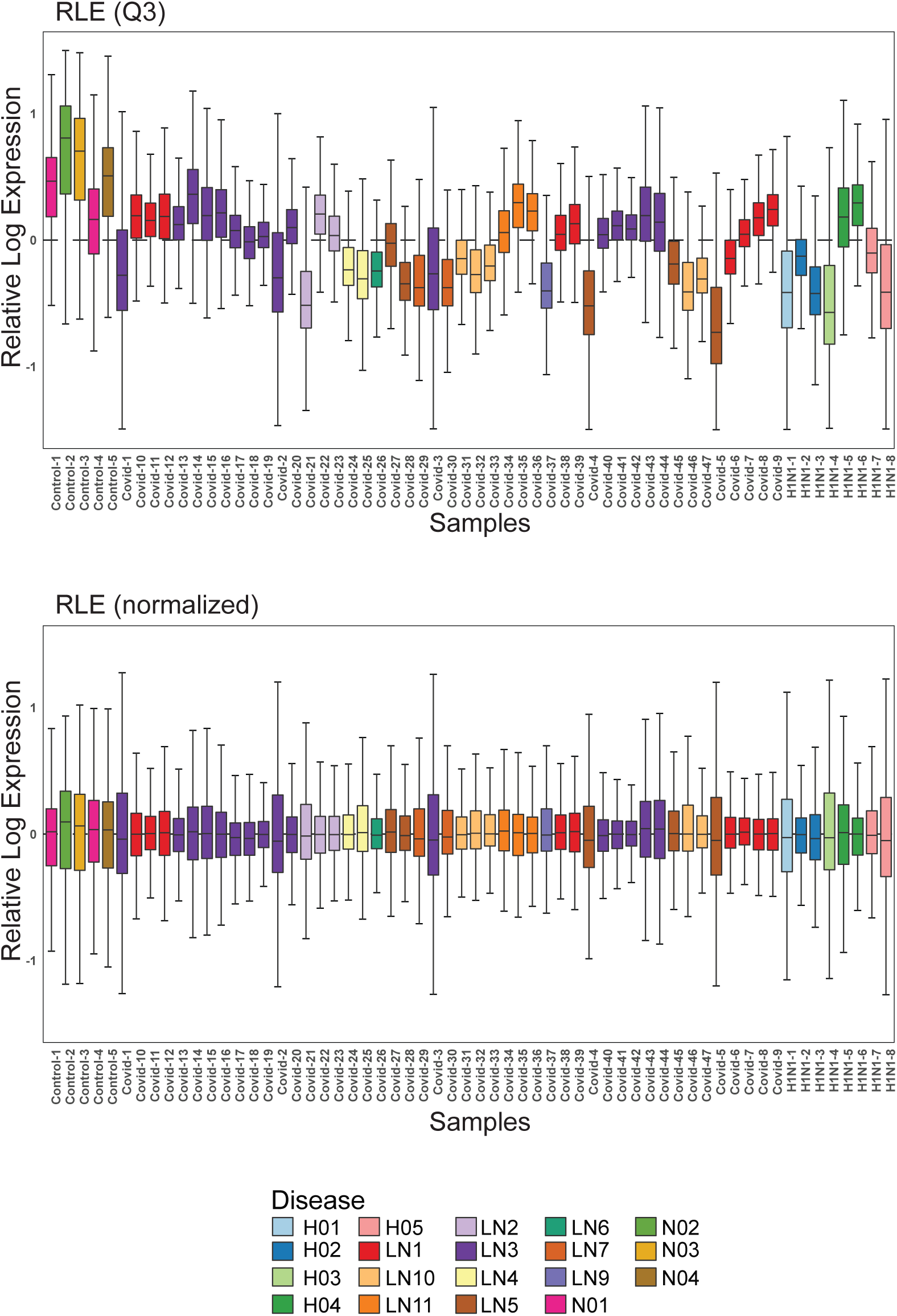
TMM normalisation of the data removed systematic bias in the uninfected samples. Relative log expression (RLE) plots for the Q3-normalised counts identified a systematic higher gene expression in the (**a**) *uninfected control* samples. The Q3-data was normalised using the (**b**) trimmed mean of M-values (TM) method^24^ using the all the genes in the gene panel. The normalisation removed the systematic bias in the regions corresponding to cores from control (uninfected) patients, allowing for cross-region analyses between the different disease types. TMM normalisation conducted using the calcNormFactors function in edgeR.

**Extended Data Figure 4.**
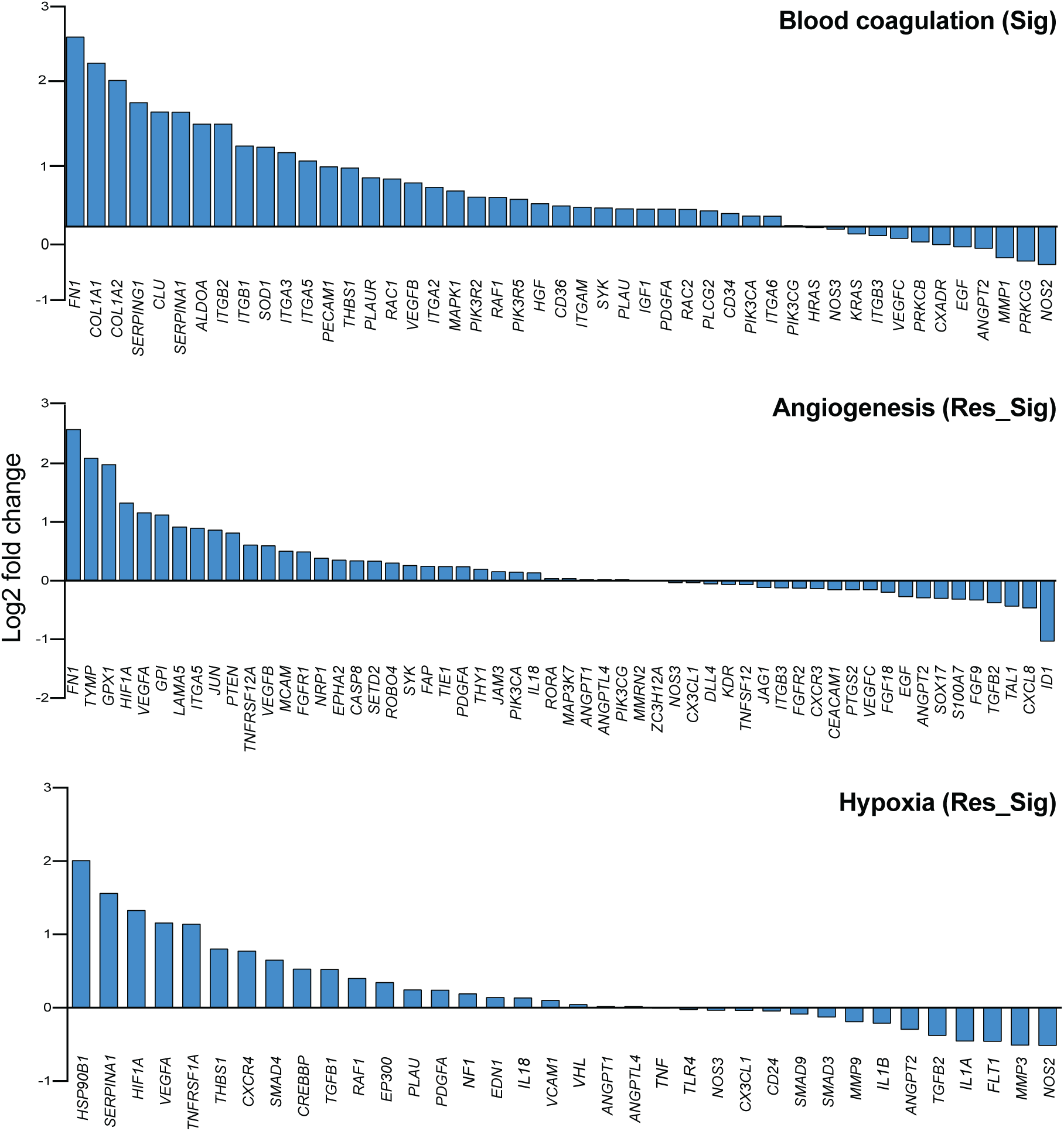

**Extended Data Figure 5.**
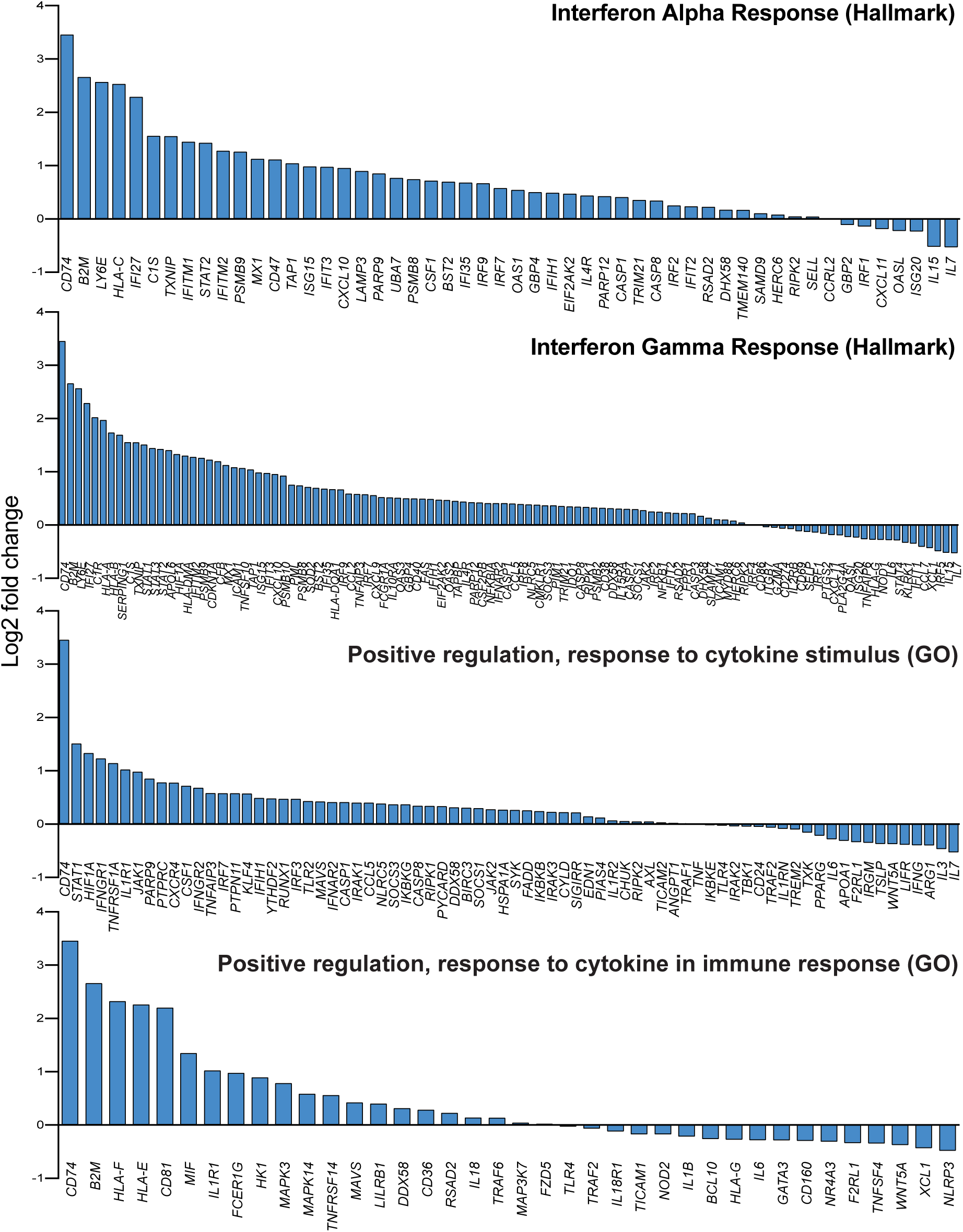

