## Extended data tables for "Spatial Profiling of Lung SARS-CoV-2 and Influenza Virus Infection Dissects Virus-Specific Host Responses and Gene Signatures"

**Extended Data Table 1**. **Patient Case History Parameters**

| **Case** | **Sex** | **Age** | **Mechanical ventilation (Days)** | **Admission to death (days)** | **Co-morbidities** | **COVID treatment** |
| --- | --- | --- | --- | --- | --- | --- |
| **LN1** | F | 85-90 | 8 | 8 | Systemic Arterial Hypertension, Dyslipidemia, Hypothyroidism, Senile dementia | Hydroxychloroquine, Azithromycin, Oseltamivir, Piperacillin, Tazobactam |
| **LN2** | M | 50-55 | 8 | 13 | Class II Obesity | Hydroxychloroquine, Azithromycin, Oseltamivir, Ceftriaxone |
| **LN3** | F | 80-85 | 0 | 23 | Systemic Arterial Hypertension, Brain Stroke, Vascular Dementia | Piperacilin, tazobactam |
| **LN4** | M | 70-75 | 10 | 38 | Type 2 Diabetes Mellitus, Chronic Kidney Disease Dialysis, Atrial Fibrillation, Coronary Disease, Heart Failure, Peripheral Obstructive Artery Disease | Hydroxychloroquine, Azithromycin, Oseltamivir, Metronidazol, Meropenen, Linezolida |
| **LN6** | M | 75-80 | 21 | 23 | Systemic Arterial Hypertension, Coronary Disease, Heart Failure, Class III obesity | Hydroxychloroquine, Azithromycin, Oseltamivir, Ceftriaxone |
| **LN7** | M | 80-85 | 8 | 8 | Systemic Arterial Hypertension, Chronic Kidney Disease Dialysis, Brain Stroke | Hydroxychloroquine, Azithromycin, Oseltamivir, Meropenen, Linezolida |
| **LN8** | F | 65-70 | 14 | 20 | Type 2 Diabetes Mellitus Systemic Arterial Hypertension  Liver transplant, Viral hepatitis,  Bladder cancer | Azithromycin, Oseltamivir, Enoxaparin, Prednisone. Piperacillin + Tazobactam |
| **LN9** | M | 85-90 | 3 | 6 | Prostate cancer, Abdominal Aortic Aneurysm, Giant Cells Arteritis | Hydroxychloroquine, Azithromycin, Oseltamivir, Meropenen |
| **LN10** | M | 45-50 | 5 | 8 | Dyslipidemia | Azithromycin, Dexamethasone |
| **LN11** | F | 90-95 | 6 | 6 | Type 2 Diabetes Mellitus, Systemic Arterial Hypertension, Dyslipidemia, Senile dementia | Azithromycin, Oseltamivir, Meropenen, Enoxaparin, Dexamethasone |

**Extended Data Table 2.** **Patient Clinical Laboratory Parameters**

| **Case** | **CRP (mg/L)** | **D-Dimer (µg/mL)** | **Leukocytes** | **Neutrophils** | **Band Cells** | **Lymphocytes** | **Hemo-globin (g/dL)** | **Platelets** | **CRP** | **D-Dimer (µg/mL** | **Leukocytes** | **Neutrophils** | **Band Cells** | **Lymphocytes** | **Hemoglobin (g/dL)** | **Platelets** |
| --- | --- | --- | --- | --- | --- | --- | --- | --- | --- | --- | --- | --- | --- | --- | --- | --- |
|  | Ref: <5mg/L | Ref: <500µg/mL | Ref: 3600 to 12000/mm³ | Ref: 1440 to 9600/ mm³ | Ref: 0 to 600/ mm³ | Ref: 720 to 5040/ mm³ | Ref: 12.5-17 g/dL | Ref: 150000 to 450000/ mm³ | Ref: <5mg/L | Ref: <500µg/ml | Ref: 3600 to 12000/mm³ | Ref: 1440 to 900/mm³ | Ref: 0 to 600/ mm³ | Ref: 720 to 5040/ mm³ | Ref:12.5-17 g/dL | Ref: 150000 to 450000/ mm³ |
|  | Initial tests |  |  |  |  |  |  |  | Final tests |  |  |  |  |  |  |  |
| **LN1** | 313 | 2014 | 15100 | 13892 | 1208 | 604 | 8.7 | 366000 | 201 | NA | 15000 | 12750 | 2400 | 1200 | 9.5 | 239000 |
| **LN2** | 146 | 425 | 11000 | 9790 | 550 | 990 | 13.5 | 325000 | 133 | 7394 | 50200 | 40160 | 3012 | 3514 | 7.2 | 318000 |
| **LN3** | 393 | NA | 19000 | 7623 | 396 | 570 | 9.9 | 282000 | 139 | NA | 9900 | 8118 | 693 | 891 | 10 | 481000 |
| **LN4** | 83 | 3436 | 9200 | 7176 | 1932 | 552 | 8.6 | 38000 | 270 | NA | 22000 | 4268 | 2420 | 440 | 8 | 356000 |
| **LN6** | 52 | 816 | 4700 | 3807 | 188 | 423 | 12.8 | 112000 | 407 | 4507 | 9400 | 7802 | 1034 | 1316 | 9.7 | 142000 |
| **LN7** | 301 | 13662 | 11800 | 11210 | 1770 | 354 | 10.7 | 252000 | 291 | NA | 24900 | 23406 | 1743 | 498 | 9.1 | 175000 |
| **LN8** | 16 | 4160 | 7380 | 5850 | N/A | 820 | 12.5 | 218000 | 16 | 1129 | 15880 | 11910 | N/A | 1260 | 7.6 | 266000 |
| **LN9** | 105 | 11184 | 8300 | 5133 | 708 | 249 | 11.5 | 110000 | 307 | 13535 | 8900 | 7921 | 445 | 712 | 9 | 99000 |
| **LN10** | 155 | 594 | 4500 | 3285 | 315 | 1035 | 15.2 | 138000 | 27.5 | 765 | 19500 | 17355 | 1365 | 975 | 10.2 | 175000 |
| **LN11** | 107 | 6544 | 7100 | 6319 | 355 | 639 | 9.5 | 111000 | 330 | N/A | 15600 | 14820 | 1092 | 780 | 8.1 | 76000 |

**Extended Data Table 3. H1N1 and Control Patient Case History Parameters**

| **Group** | **Case** | **Cause of death** | **Sex** | **Age** | **Mechanical ventilation (Days)** | **Days from admission**  **to death** | **Comorbidities** |
| --- | --- | --- | --- | --- | --- | --- | --- |
| **H1N1** | 99-6999 | H1N1 | M | 50-55 | 19 | 19 | Haemophilus influenza |
| **H1N1** | 99-7000 | H1N1 | M | 55-60 | 12 | 12 | Haemophilus influenza |
| **H1N1** | 99-7223 | H1N1 | M | 30-35 | 1 | 1 | Mycoplasma pneumoniae |
| **H1N1** | 99-7888 | H1N1 | M | 35-40 | 1 | 1 | Haemophilus influenza |
| **H1N1** | 99-8157 | H1N1 | M | 50-55 | 3 | 3 | None |
| **Control** | N07-09 | Neuroendocrine CA | M | 40-45 |  |  |  |
| **Control** | N08-21 | Hepatic CA | M | 55-60 |  |  |  |
| **Control** | N09-19 | Surgery | M | 20-25 |  |  |  |
| **Control** | N09-57 | Lymphoma | M | 20-25 |  |  |  |

**Extended Data Table 4.** **Histopathological Parameters**

| **Group** | **Case** | **Fibrosis** | **Histology** |
| --- | --- | --- | --- |
| **COVID** | LN1 | Anatomical fibrosis | Acute phase DAD |
| **COVID** | LN2 | Anatomical fibrosis | Acute phase DAD |
| **COVID** | LN3 | Anatomical fibrosis | Acute phase DAD |
| **COVID** | LN4 | Anatomical fibrosis | Reactive Type 2 pneumocyte hyperplasia |
| **COVID** | LN6 | Anatomical fibrosis | Reactive Type 2 pneumocyte hyperplasia |
| **COVID** | LN7 | Anatomical fibrosis | Reactive Type 2 pneumocyte hyperplasia |
| **COVID** | LN8 | Alveolar, moderate interstitial fibrosis | Organising pneumonia |
| **COVID** | LN9 | Anatomical fibrosis | Organising pneumonia |
| **COVID** | LN10 | Anatomical fibrosis | Acute phase DAD |
| **COVID** | LN11 | Anatomical fibrosis | Acute phase DAD |
| **H1N1** | 99-6999 | Anatomical fibrosis | Alveolar haemorrhage |
| **H1N1** | 99-7000 | Anatomical fibrosis | Alveolar haemorrhage |
| **H1N1** | 99-7223 | Anatomical fibrosis | Type 2 pneumocyte hyperplasia |
| **H1N1** | 99-7888 | Mild interstitial fibrosis | Acute phase DAD |
| **H1N1** | 99-8157 | Mild interstitial fibrosis | Late organising phase DAD |
| **Control** | N07-09 | Anatomical fibrosis | Nil |
| **Control** | N08-21 | Anatomical fibrosis | Carcinoma primary or secondary |
| **Control** | N09-19 | Anatomical fibrosis | Pulmonary oedema |
| **Control** | N09-57 | Anatomical fibrosis | Pulmonary oedema |

* Histology of FFPE cores was assessed by pathologist for levels of fibrosis, organisation and tissue injury. DAD; Diffuse Alveolar Damage

**Extended Data Table 5.** **Histopathological Scoring of ROIs**

| **ROI#** | **Group** | **Case** | **Dominant Tissue Type** | **H&E** | **H&E** | **H&E** | **H&E** | **H&E** | **H&E** | **Spike IHC** | **Spike IHC** | **Spike IHC** | **Spike IHC** | **RNAscope Score** |
| --- | --- | --- | --- | --- | --- | --- | --- | --- | --- | --- | --- | --- | --- | --- |
|  |  |  |  | Alveolar Haemorrhage | Hyaline Membranes | Type 2 Pneumocyte Hyperplasia | Capillary Congestion | Fibroblastic Foci | Interstitial Inflammation | Bronchiolar Epithelium | Type 2 Pneumocytes | Interstitial Lymphocytes | Alveolar Macrophages |  |
| **10** | COVID | LN1 | Hyaline membrane | 1 | 3 | 2 | 2 | 0 | 0 | 0 | 1 | 0 | 0 | 1 |
| **11** | COVID | LN1 | Hyaline membrane | 0 | 3 | 3 | 3 | 0 | 0 | 0 | 2 | 0 | 3 | 1 |
| **12** | COVID | LN1 | Hyaline membrane | 1 | 3 | 3 | 3 | 0 | 0 | 0 | 2 | 0 | 0 | 1 |
| **6** | COVID | LN1 | Hyaline membranes | 0 | 3 | 3 | 3 | 0 | 0 | 0 | 2 | 0 | 3 | 3 |
| **7** | COVID | LN1 | Hyaline membranes, type 2 pneumocytes | 0 | 3 | 3 | 2 | 0 | 0 | 0 | 2 | 1 | 3 | 3 |
| **8** | COVID | LN1 | Hyaline membranes, type 2 pneumocytes | 0 | 3 | 3 | 2 | 0 | 0 | 0 | 2 | 0 | 3 | 2 |
| **9** | COVID | LN1 | Hyaline membranes, type 2 pneumocytes | 1 | 3 | 2 | 2 | 0 | 0 | 0 | 0 | 0 | 0 | 1 |
| **38** | COVID | LN1 | Type 2 pneumocytes | 1 | 0 | 3 | 3 | 0 | 0 | 0 | 2 | 0 | 3 | 2 |
| **39** | COVID | LN1 | Type 2 pneumocytes | 1 | 3 | 2 | 2 | 0 | 0 | 0 | 2 | 0 | 0 | 1 |
| **22** | COVID | LN2 | Lymphocytes | 0 | 0 | 0 | 0 | 0 | 3 | 0 | 0 | 2 | 0 | 0 |
| **23** | COVID | LN2 | Lymphocytes and type 2 pneumocytes | 0 | 0 | 2 | 2 | 0 | 2 | 0 | 2 | 2 | 0 | 0 |
| **21** | COVID | LN2 | Type 2 pneumocytes | 0 | 0 | 3 | 3 | 0 | 0 | 0 | 2 | 0 | 0 | 0 |
| **1** | COVID | LN3 | Bronchiolar epithelium | 0 | 0 | 0 | 0 | 0 | 0 | 3 | 0 | 0 | 0 | 0 |
| **2** | COVID | LN3 | Bronchiolar epithelium | 0 | 0 | 0 | 0 | 0 | 0 | 3 | 0 | 0 | 0 | 0 |
| **3** | COVID | LN3 | Bronchiolar epithelium | 0 | 0 | 0 | 0 | 0 | 0 | 3 | 0 | 0 | 0 | 0 |
| **14** | COVID | LN3 | Hyaline membranes | 0 | 3 | 0 | 0 | 0 | 0 | 0 | 1 | 1 | 3 | 2 |
| **13** | COVID | LN3 | Macrophages | 0 | 3 | 2 | 3 | 0 | 1 | 0 | 0 | 1 | 3 | 1 |
| **15** | COVID | LN3 | Macrophages | 2 | 3 | 2 | 3 | 0 | 2 | 0 | 2 | 1 | 2 | 1 |
| **16** | COVID | LN3 | Macrophages | 0 | 3 | 0 | 3 | 0 | 0 | 0 | 0 | 0 | 3 | 1 |
| **17** | COVID | LN3 | Macrophages | 0 | 0 | 3 | 3 | 0 | 0 | 0 | 2 | 0 | 3 | 2 |
| **19** | COVID | LN3 | Macrophages | 0 | 3 | 3 | 3 | 0 | 1 | 0 | 2 | 1 | 2 | 1 |
| **20** | COVID | LN3 | Macrophages | 0 | 3 | 2 | 3 | 0 | 0 | 0 | 1 | 1 | 3 | 1 |
| **40** | COVID | LN3 | Macrophages | 0 | 3 | 0 | 3 | 0 | 1 | 0 | 2 | 1 | 2 | 0 |
| **42** | COVID | LN3 | Macrophages | 0 | 3 | 2 | 3 | 0 | 0 | 0 | 2 | 1 | 3 | 1 |
| **43** | COVID | LN3 | Subepithelial lymphocytes | 0 | 0 | 0 | 3 | 0 | 1 | 0 | 0 | 1 | 0 | 0 |
| **44** | COVID | LN3 | Subepithelial lymphocytes | 0 | 0 | 0 | 3 | 0 | 1 | 0 | 0 | 2 | 0 | 0 |
| **18** | COVID | LN3 | Type 2 pneumocytes | 0 | 0 | 2 | 3 | 0 | 1 | 0 | 2 | 1 | 3 | 2 |
| **41** | COVID | LN3 | Type 2 pneumocytes | 0 | 3 | 2 | 3 | 0 | 1 | 0 | 2 | 2 | 3 | 0 |
| **24** | COVID | LN4 | Type 2 pneumocytes | 0 | 0 | 3 | 3 | 0 | 0 | 0 | 3 | 0 | 0 | 0 |
| **25** | COVID | LN4 | Type 2 pneumocytes | 0 | 0 | 3 | 3 | 0 | 0 | 0 | 3 | 0 | 3 | 0 |
| **4** | COVID | LN6 | Bronchiolar epithelium | 0 | 0 | 0 | 0 | 0 | 0 | 3 | 0 | 0 | 0 | 0 |
| **5** | COVID | LN6 | Bronchiolar epithelium | 0 | 0 | 0 | 0 | 0 | 0 | 3 | 0 | 0 | 0 | 0 |
| **27** | COVID | LN6 | Type 2 pneumocytes | 0 | 0 | 1 | 0 | 0 | 1 | 0 | 1 | 1 | 0 | 0 |
| **28** | COVID | LN6 | Type 2 pneumocytes | 0 | 0 | 2 | 2 | 0 | 0 | 0 | 2 | 0 | 0 | 0 |
| **45** | COVID | LN6 | Type 2 pneumocytes | 0 | 0 | 3 | 2 | 0 | 1 | 0 | 1 | 1 | 0 | 0 |
| **26** | COVID | LN7 | Type 2 pneumocytes | 0 | 0 | 1 | 2 | 0 | 1 | 0 | 2 | 2 | 0 | 0 |
| **29** | COVID | LN8 | Macrophages | 3 | 0 | 3 | 3 | 3 | 3 | 0 | 2 | 2 | 3 | 0 |
| **30** | COVID | LN8 | Macrophages | 3 | 0 | 3 | 3 | 3 | 3 | 0 | 1 | 2 | 3 | 0 |
| **37** | COVID | LN9 | Macrophages | 1 | 0 | 2 | 3 | 3 | 2 | 0 | 2 | 2 | 3 | 0 |
| **31** | COVID | LN10 | Type 2 pneumocytes and Bronchiolar epithelium | 0 | 0 | 2 | 3 | 0 | 3 | 3 | 2 | 2 | 2 | 0 |
| **32** | COVID | LN10 | Type 2 pneumocytes | 2 | 0 | 3 | 3 | 1 | 2 | 0 | 3 | 2 | 2 | 0 |
| **33** | COVID | LN10 | Type 2 pneumocytes | 1 | 3 | 3 | 3 | 0 | 0 | 0 | 3 | 1 | 3 | 0 |
| **46** | COVID | LN10 | Type 2 pneumocytes | 1 | 1 | 3 | 1 | 0 | 1 | 0 | 3 | 2 | 2 | 0 |
| **47** | COVID | LN10 | Type 2 pneumocytes | 1 | 0 | 3 | 2 | 0 | 1 | 0 | 3 | 2 | 3 | 0 |
| **34** | COVID | LN11 | Bronchiolar epithelium | 0 | 0 | 0 | 0 | 0 | 0 | 3 | 0 | 0 | 0 | 0 |
| **36** | COVID | LN11 | Hyaline membranes | 0 | 3 | 2 | 0 | 0 | 2 | 0 | 3 | 2 | 2 | 0 |
| **35** | COVID | LN11 | Type 2 pneumocytes | 0 | 3 | 3 | 0 | 0 | 1 | 0 | 2 | 2 | 3 | 0 |
| **8** | H1N1 | N09-6999 | Bronchiolar epithelium | 0 | 0 | 0 | 0 | 0 | 0 | 3 | 0 | 0 | 0 |  |
| **7** | H1N1 | N09-6999 | Macrophages | 3 | 0 | 3 | 0 | 0 | 3 | 0 | 2 | 0 | 1 |  |
| **1** | H1N1 | N09-7000 | Bronchiolar epithelium | 0 | 0 | 0 | 0 | 0 | 0 | 2 | 0 | 0 | 0 |  |
| **2** | H1N1 | N09-7223 | Macrophages | 2 | 0 | 3 | 3 | 0 | 0 | 0 | 0 | 0 | 1 |  |
| **3** | H1N1 | N09-7223 | Macrophages | 2 | 0 | 3 | 3 | 0 | 0 | 0 | 1 | 0 | 2 |  |
| **5** | H1N1 | N09-7888 | Intravascular lymphocytes | 0 | 0 | 0 | 0 | 0 | 0 | 0 | 0 | 0 | 0 |  |
| **6** | H1N1 | N09-7888 | Type 2 pneumocytes | 1 | 3 | 3 | 3 | 0 | 0 | 0 | 0 | 0 | 1 |  |
| **4** | H1N1 | N09-8157 | Type 2 pneumocytes | 1 | 0 | 2 | 0 | 0 | 3 | 0 | 1 | 1 | 0 |  |
| **5** | NORMAL | N07-009 | Type 2 pneumocytes | 1 | 0 | 0 | 3 | 0 | 0 | 0 | 0 | 0 | 0 |  |
| **3** | NORMAL | N08-021 | Carcinoma | 0 | 0 | 0 | 0 | 0 | 0 | 0 | 0 | 0 | 0 |  |
| **4** | NORMAL | N09-019 | Bronchiolar epithelium | 0 | 0 | 0 | 0 | 0 | 0 | 2 | 0 | 0 | 1 |  |
| **1** | NORMAL | N09-019 | Macrophages | 0 | 0 | 1 | 0 | 0 | 0 | 0 | 0 | 0 | 0 |  |
| **2** | NORMAL | N09-57 | Macrophages | 0 | 0 | 1 | 0 | 0 | 0 | 0 | 0 | 0 | 0 |  |

**Extended Data Table 6.** **Genes downregulated in the lungs of SARS-CoV-2 patients compared with non-virally infected controls.**

| **Gene name** | **Fold Change (log2)** | **Average Expression** | **t-statistics** | **p-value** | **Adjusted p-value** |
| --- | --- | --- | --- | --- | --- |
| CSF3 | -1.060 | 6.642 | -2.740 | 0.00401 | 0.02794 |
| PPP2R1A | -0.817 | 7.657 | -3.234 | 0.00098 | 0.00819 |
| IFNA8 | -0.728 | 7.554 | -2.695 | 0.00452 | 0.03075 |
| MAGEC2 | -0.704 | 7.792 | -3.019 | 0.00184 | 0.01440 |
| PCK1 | -0.694 | 7.777 | -3.596 | 0.00032 | 0.00313 |
| SARS-CoV-2-Neg | -0.683 | 7.714 | -2.842 | 0.00302 | 0.02212 |
| NegProbe | -0.672 | 7.483 | -4.324 | 0.00003 | 0.00035 |
| MLANA | -0.658 | 7.558 | -3.325 | 0.00074 | 0.00639 |
| NAALAD2 | -0.596 | 7.650 | -2.722 | 0.00420 | 0.02883 |
| CD19 | -0.593 | 7.831 | -2.685 | 0.00464 | 0.03149 |
| FGF17 | -0.582 | 7.318 | -2.496 | 0.00761 | 0.04729 |
| MPL | -0.582 | 7.875 | -2.810 | 0.00330 | 0.02351 |
| IL9 | -0.575 | 7.721 | -2.662 | 0.00493 | 0.03323 |
| GAGE1 | -0.567 | 7.469 | -2.651 | 0.00507 | 0.03380 |
| IL13 | -0.567 | 7.716 | -2.488 | 0.00778 | 0.04784 |
| PRAME | -0.561 | 7.564 | -2.517 | 0.00721 | 0.04529 |
| IFNL1 | -0.558 | 7.762 | -2.473 | 0.00808 | 0.04905 |
| LDHA | -0.542 | 7.791 | -2.479 | 0.00794 | 0.04852 |
| C9 | -0.530 | 7.670 | -2.511 | 0.00733 | 0.04586 |
| IL7 | -0.526 | 7.794 | -2.553 | 0.00656 | 0.04192 |

**Extended Data Table 7.** **Genes upregulated in the lungs of SARS-CoV-2 patients compared with non-virally infected controls.**

| **Gene name** | **Fold Change (log2)** | **Average Expression** | **t-statistics** | **p-value** | **Adjusted p-value** |
| --- | --- | --- | --- | --- | --- |
| CD74 | 3.456 | 13.824 | 10.579 | 7.54E-16 | 2.79E-13 |
| SSX1 | 3.026 | 11.624 | 17.734 | 2.38E-26 | 4.41E-23 |
| COL3A1 | 2.822 | 11.546 | 5.737 | 1.53E-07 | 3.83E-06 |
| B2M | 2.660 | 14.118 | 8.550 | 2.11E-12 | 1.78E-10 |
| HLA-DRA | 2.659 | 11.577 | 9.218 | 1.49E-13 | 1.85E-11 |
| ACTB | 2.612 | 14.090 | 7.752 | 5.15E-11 | 2.89E-09 |
| FN1 | 2.587 | 12.345 | 6.410 | 1.10E-08 | 3.23E-07 |
| **LY6E** | 2.571 | 10.949 | 11.983 | 3.96E-18 | 3.67E-15 |
| SFTPA1 | 2.563 | 13.247 | 2.945 | 2.57E-03 | 1.91E-02 |
| HLA-C | 2.532 | 12.090 | 10.324 | 2.00E-15 | 6.16E-13 |
| HLA-F | 2.323 | 11.219 | 10.984 | 1.62E-16 | 7.48E-14 |
| CD63 | 2.298 | 11.385 | 9.479 | 5.33E-14 | 8.97E-12 |
| **IFI27** | 2.289 | 10.932 | 7.896 | 2.88E-11 | 1.72E-09 |
| ANXA1 | 2.280 | 10.400 | 11.337 | 4.30E-17 | 2.65E-14 |
| HLA-E | 2.262 | 11.640 | 8.574 | 1.92E-12 | 1.69E-10 |
| KRT7 | 2.259 | 10.763 | 4.324 | 2.85E-05 | 3.52E-04 |
| COL1A1 | 2.234 | 11.022 | 4.423 | 2.02E-05 | 2.59E-04 |
| CD163 | 2.216 | 10.854 | 7.565 | 1.09E-10 | 5.64E-09 |
| CD81 | 2.203 | 11.362 | 8.971 | 3.95E-13 | 4.30E-11 |
| FLNA | 2.186 | 10.894 | 7.563 | 1.10E-10 | 5.64E-09 |
| KRT18 | 2.133 | 11.108 | 5.014 | 2.36E-06 | 3.87E-05 |
| CD68 | 2.127 | 10.776 | 9.258 | 1.27E-13 | 1.82E-11 |
| TYMP | 2.093 | 10.862 | 8.715 | 1.09E-12 | 1.07E-10 |
| C1QA | 2.087 | 10.690 | 8.302 | 5.68E-12 | 4.05E-10 |
| KRT19 | 2.055 | 9.979 | 4.589 | 1.11E-05 | 1.56E-04 |
| A2M | 2.038 | 12.558 | 5.212 | 1.13E-06 | 2.04E-05 |
| C1R | 2.025 | 10.440 | 7.179 | 5.11E-10 | 2.15E-08 |
| APP | 2.017 | 10.607 | 9.011 | 3.38E-13 | 3.91E-11 |
| HSP90B1 | 2.012 | 10.113 | 9.217 | 1.50E-13 | 1.85E-11 |
| CTSS | 2.008 | 10.175 | 9.804 | 1.50E-14 | 3.46E-12 |
| COL1A2 | 1.998 | 11.096 | 4.787 | 5.43E-06 | 8.04E-05 |
| GPX1 | 1.985 | 11.060 | 8.944 | 4.41E-13 | 4.54E-11 |
| HLA-A | 1.975 | 12.878 | 7.191 | 4.87E-10 | 2.10E-08 |
| OAZ1 | 1.946 | 11.412 | 8.360 | 4.50E-12 | 3.47E-10 |
| CTSH | 1.925 | 10.501 | 4.983 | 2.64E-06 | 4.24E-05 |
| CD9 | 1.905 | 10.245 | 6.956 | 1.25E-09 | 4.71E-08 |
| LGALS3 | 1.861 | 10.319 | 8.577 | 1.89E-12 | 1.69E-10 |
| CCL18 | 1.857 | 10.005 | 5.360 | 6.42E-07 | 1.28E-05 |
| UBB | 1.856 | 11.016 | 8.325 | 5.19E-12 | 3.84E-10 |
| DDIT4 | 1.853 | 10.493 | 5.811 | 1.14E-07 | 2.89E-06 |
| HLA-DRB3 | 1.833 | 10.665 | 6.021 | 5.04E-08 | 1.39E-06 |
| LAMP1 | 1.781 | 10.410 | 9.314 | 1.02E-13 | 1.57E-11 |
| CTNNB1 | 1.770 | 9.983 | 9.668 | 2.54E-14 | 5.23E-12 |
| LYZ | 1.764 | 10.533 | 5.294 | 8.25E-07 | 1.57E-05 |
| HLA-B | 1.735 | 13.623 | 5.849 | 9.83E-08 | 2.57E-06 |
| H3C10 | 1.734 | 10.187 | 5.658 | 2.06E-07 | 4.92E-06 |
| HLA-DRB4 | 1.727 | 10.644 | 4.854 | 4.23E-06 | 6.42E-05 |
| PKM | 1.716 | 10.692 | 9.615 | 3.13E-14 | 5.80E-12 |
| SERPING1 | 1.693 | 11.532 | 4.841 | 4.43E-06 | 6.67E-05 |
| GNAS | 1.686 | 11.510 | 5.717 | 1.64E-07 | 4.05E-06 |
| RPL7A | 1.679 | 11.626 | 5.656 | 2.07E-07 | 4.92E-06 |
| CAPN2 | 1.663 | 10.033 | 6.528 | 6.84E-09 | 2.11E-07 |
| CD14 | 1.658 | 9.507 | 6.540 | 6.53E-09 | 2.08E-07 |
| COL5A1 | 1.637 | 9.452 | 4.485 | 1.60E-05 | 2.13E-04 |
| COL6A3 | 1.628 | 10.820 | 4.432 | 1.93E-05 | 2.53E-04 |
| CEACAM6 | 1.623 | 10.178 | 3.871 | 1.32E-04 | 1.45E-03 |
| FCGRT | 1.613 | 9.893 | 8.366 | 4.40E-12 | 3.47E-10 |
| ADH1A | 1.609 | 9.753 | 5.614 | 2.44E-07 | 5.41E-06 |
| C3 | 1.589 | 10.763 | 3.336 | 7.27E-04 | 6.35E-03 |
| CLU | 1.567 | 10.458 | 3.590 | 3.28E-04 | 3.20E-03 |
| SERPINA1 | 1.564 | 10.750 | 3.803 | 1.65E-04 | 1.76E-03 |
| C1S | 1.556 | 9.915 | 5.640 | 2.20E-07 | 5.17E-06 |
| TXNIP | 1.553 | 10.982 | 4.240 | 3.76E-05 | 4.53E-04 |
| CD44 | 1.539 | 10.424 | 6.465 | 8.77E-09 | 2.66E-07 |
| RPS27A | 1.537 | 10.665 | 5.344 | 6.82E-07 | 1.33E-05 |
| CCND1 | 1.520 | 9.444 | 4.602 | 1.05E-05 | 1.50E-04 |
| ACTA2 | 1.511 | 9.385 | 3.619 | 2.98E-04 | 2.98E-03 |
| STAT1 | 1.510 | 9.820 | 6.549 | 6.31E-09 | 2.08E-07 |
| C1QB | 1.498 | 9.642 | 7.111 | 6.70E-10 | 2.76E-08 |
| RAB7A | 1.482 | 9.890 | 10.081 | 5.11E-15 | 1.35E-12 |
| **IFI6** | 1.476 | 9.913 | 5.091 | 1.77E-06 | 3.06E-05 |
| IFITM1 | 1.447 | 10.502 | 5.016 | 2.33E-06 | 3.86E-05 |
| STAT3 | 1.444 | 10.017 | 6.934 | 1.36E-09 | 5.04E-08 |
| MUC1 | 1.437 | 10.106 | 3.188 | 1.13E-03 | 9.35E-03 |
| BAX | 1.430 | 9.060 | 8.024 | 1.73E-11 | 1.07E-09 |
| RPS6 | 1.428 | 11.502 | 3.915 | 1.13E-04 | 1.27E-03 |
| STAT2 | 1.427 | 9.587 | 8.199 | 8.58E-12 | 5.67E-10 |
| LRP1 | 1.421 | 10.286 | 6.209 | 2.41E-08 | 6.86E-07 |
| C7 | 1.411 | 9.782 | 3.621 | 2.96E-04 | 2.98E-03 |
| APOL6 | 1.406 | 9.456 | 6.537 | 6.60E-09 | 2.08E-07 |
| ALDOA | 1.405 | 11.133 | 5.569 | 2.89E-07 | 6.15E-06 |
| ITGB2 | 1.404 | 9.841 | 5.884 | 8.59E-08 | 2.27E-06 |
| TPSB2 | 1.382 | 9.653 | 3.640 | 2.78E-04 | 2.81E-03 |
| CD97 | 1.360 | 9.880 | 7.046 | 8.71E-10 | 3.43E-08 |
| MIF | 1.348 | 9.717 | 6.914 | 1.47E-09 | 5.35E-08 |
| HIF1A | 1.331 | 9.341 | 7.236 | 4.06E-10 | 1.84E-08 |
| COX6A1 | 1.326 | 10.440 | 6.442 | 9.62E-09 | 2.87E-07 |
| BRD2 | 1.324 | 9.546 | 8.252 | 6.94E-12 | 4.76E-10 |
| CD164 | 1.316 | 9.799 | 7.197 | 4.75E-10 | 2.09E-08 |
| CTSL | 1.315 | 10.252 | 4.860 | 4.14E-06 | 6.34E-05 |
| H3-3A | 1.310 | 10.042 | 6.200 | 2.50E-08 | 7.01E-07 |
| CD55 | 1.302 | 10.089 | 3.305 | 7.92E-04 | 6.76E-03 |
| HLA-DMA | 1.302 | 9.545 | 5.506 | 3.69E-07 | 7.58E-06 |
| SF3B1 | 1.301 | 9.651 | 7.236 | 4.07E-10 | 1.84E-08 |
| NDUFA13 | 1.301 | 10.040 | 7.072 | 7.84E-10 | 3.16E-08 |
| TPM4 | 1.297 | 10.618 | 4.747 | 6.25E-06 | 9.18E-05 |
| LDHB | 1.295 | 9.866 | 4.482 | 1.61E-05 | 2.14E-04 |
| JUNB | 1.291 | 10.170 | 4.907 | 3.48E-06 | 5.50E-05 |
| CD99 | 1.288 | 9.679 | 7.587 | 9.99E-11 | 5.44E-09 |
| CD4 | 1.281 | 9.295 | 7.296 | 3.20E-10 | 1.56E-08 |
| EWSR1 | 1.279 | 10.008 | 7.851 | 3.46E-11 | 2.00E-09 |
| IFITM2 | 1.278 | 10.807 | 4.428 | 1.96E-05 | 2.55E-04 |
| MDM2 | 1.275 | 9.172 | 5.665 | 2.00E-07 | 4.88E-06 |
| DUSP1 | 1.268 | 10.724 | 3.429 | 5.40E-04 | 5.02E-03 |
| H3C2 | 1.267 | 8.791 | 3.406 | 5.80E-04 | 5.34E-03 |
| PSMB9 | 1.260 | 9.919 | 5.612 | 2.46E-07 | 5.41E-06 |
| CXCL16 | 1.253 | 9.219 | 5.272 | 8.93E-07 | 1.67E-05 |
| SLC1A5 | 1.241 | 9.260 | 5.592 | 2.65E-07 | 5.78E-06 |
| ENG | 1.239 | 10.026 | 4.293 | 3.13E-05 | 3.79E-04 |
| HLA-DPA1 | 1.234 | 9.398 | 6.573 | 5.73E-09 | 1.93E-07 |
| IFNGR1 | 1.232 | 9.428 | 6.229 | 2.23E-08 | 6.45E-07 |
| IDH1 | 1.232 | 9.349 | 5.575 | 2.83E-07 | 6.10E-06 |
| GRB2 | 1.229 | 9.422 | 8.060 | 1.49E-11 | 9.54E-10 |
| CDKN1A | 1.226 | 9.848 | 4.181 | 4.62E-05 | 5.52E-04 |
| H3-5 | 1.226 | 9.942 | 5.021 | 2.29E-06 | 3.85E-05 |
| SREBF1 | 1.224 | 9.546 | 4.050 | 7.19E-05 | 8.33E-04 |
| PRDX5 | 1.201 | 9.811 | 4.870 | 3.99E-06 | 6.19E-05 |
| CFB | 1.199 | 9.777 | 3.790 | 1.71E-04 | 1.82E-03 |
| COX5B | 1.194 | 9.909 | 7.000 | 1.05E-09 | 4.04E-08 |
| VEGFA | 1.163 | 9.514 | 3.966 | 9.57E-05 | 1.09E-03 |
| NDUFB4 | 1.156 | 9.611 | 7.478 | 1.54E-10 | 7.72E-09 |
| GDF15 | 1.156 | 8.995 | 3.007 | 1.91E-03 | 1.47E-02 |
| BCL2L1 | 1.145 | 9.540 | 4.368 | 2.41E-05 | 3.04E-04 |
| TNFRSF1A | 1.145 | 9.966 | 5.291 | 8.33E-07 | 1.57E-05 |
| CD46 | 1.143 | 9.585 | 4.053 | 7.12E-05 | 8.29E-04 |
| AKT1 | 1.141 | 9.673 | 4.991 | 2.55E-06 | 4.15E-05 |
| VCAN | 1.138 | 9.483 | 3.485 | 4.53E-04 | 4.28E-03 |
| TNC | 1.138 | 9.525 | 2.639 | 5.29E-03 | 3.51E-02 |
| CEBPB | 1.131 | 9.756 | 5.357 | 6.49E-07 | 1.28E-05 |
| GPI | 1.129 | 9.164 | 6.537 | 6.61E-09 | 2.08E-07 |
| RHOB | 1.128 | 10.349 | 2.769 | 3.71E-03 | 2.60E-02 |
| MX1 | 1.126 | 9.748 | 2.983 | 2.04E-03 | 1.55E-02 |
| ITGAV | 1.116 | 9.074 | 5.240 | 1.01E-06 | 1.85E-05 |
| CD47 | 1.114 | 9.427 | 4.416 | 2.04E-05 | 2.61E-04 |
| ITGB1 | 1.104 | 10.627 | 3.489 | 4.48E-04 | 4.25E-03 |
| TPM1 | 1.102 | 9.776 | 2.983 | 2.04E-03 | 1.55E-02 |
| MAP2K2 | 1.101 | 9.453 | 6.643 | 4.34E-09 | 1.49E-07 |
| SOD1 | 1.087 | 10.075 | 4.533 | 1.35E-05 | 1.83E-04 |
| S100A9 | 1.085 | 11.062 | 2.568 | 6.37E-03 | 4.09E-02 |
| POLR2A | 1.075 | 9.117 | 7.245 | 3.93E-10 | 1.84E-08 |
| STAT6 | 1.074 | 9.514 | 6.699 | 3.48E-09 | 1.24E-07 |
| TNFSF10 | 1.071 | 9.246 | 3.820 | 1.55E-04 | 1.68E-03 |
| NDUFA11 | 1.069 | 9.594 | 5.562 | 2.98E-07 | 6.27E-06 |
| COL5A2 | 1.064 | 8.843 | 3.013 | 1.87E-03 | 1.45E-02 |
| MCL1 | 1.060 | 10.445 | 3.783 | 1.75E-04 | 1.85E-03 |
| ANP32B | 1.056 | 9.170 | 5.163 | 1.35E-06 | 2.42E-05 |
| ADH1B | 1.052 | 9.578 | 2.629 | 5.40E-03 | 3.57E-02 |
| CYBB | 1.050 | 8.950 | 4.316 | 2.89E-05 | 3.54E-04 |
| TAP1 | 1.045 | 10.082 | 3.427 | 5.43E-04 | 5.03E-03 |
| DAB2 | 1.044 | 9.054 | 4.704 | 7.31E-06 | 1.05E-04 |
| ILF3 | 1.043 | 9.518 | 5.512 | 3.60E-07 | 7.49E-06 |
| NDUFS8 | 1.039 | 8.971 | 5.898 | 8.14E-08 | 2.18E-06 |
| MRC1 | 1.037 | 9.450 | 2.894 | 2.62E-03 | 1.94E-02 |
| SBNO2 | 1.034 | 9.398 | 5.367 | 6.26E-07 | 1.26E-05 |
| IL6ST | 1.029 | 9.659 | 4.523 | 1.40E-05 | 1.89E-04 |
| IL1R1 | 1.024 | 9.869 | 3.332 | 7.27E-04 | 6.35E-03 |
| PDGFRB | 1.020 | 9.320 | 3.964 | 9.62E-05 | 1.09E-03 |
| TUBB | 1.020 | 9.530 | 3.945 | 1.02E-04 | 1.15E-03 |
| ATP5F1D | 1.015 | 9.477 | 5.317 | 7.56E-07 | 1.46E-05 |
| ITGA3 | 1.014 | 9.674 | 2.578 | 6.19E-03 | 4.02E-02 |
| SFN | 1.012 | 9.517 | 2.468 | 8.22E-03 | 4.97E-02 |
| CSF1R | 1.005 | 9.096 | 3.566 | 3.51E-04 | 3.40E-03 |
| COX6B1 | 0.990 | 9.843 | 4.581 | 1.13E-05 | 1.58E-04 |
| CDKN1B | 0.988 | 9.159 | 5.503 | 3.73E-07 | 7.59E-06 |
| TECR | 0.988 | 9.326 | 5.257 | 9.47E-07 | 1.75E-05 |
| ALCAM | 0.985 | 9.264 | 3.354 | 6.79E-04 | 6.08E-03 |
| JAK1 | 0.983 | 9.550 | 5.616 | 2.42E-07 | 5.41E-06 |
| NDUFB7 | 0.982 | 11.000 | 4.080 | 6.50E-05 | 7.62E-04 |
| CD276 | 0.979 | 8.998 | 5.952 | 6.60E-08 | 1.80E-06 |
| FCER1G | 0.977 | 9.521 | 3.459 | 4.92E-04 | 4.63E-03 |
| ADH1C | 0.972 | 9.747 | 3.024 | 1.81E-03 | 1.43E-02 |
| MLPH | 0.968 | 8.818 | 2.566 | 6.37E-03 | 4.09E-02 |
| SDHA | 0.961 | 8.997 | 5.613 | 2.45E-07 | 5.41E-06 |
| G6PD | 0.948 | 9.144 | 5.156 | 1.38E-06 | 2.46E-05 |
| HLA-DMB | 0.942 | 9.205 | 3.565 | 3.52E-04 | 3.40E-03 |
| CDC25B | 0.941 | 9.435 | 2.949 | 2.24E-03 | 1.68E-02 |
| ETS2 | 0.937 | 10.148 | 2.776 | 3.63E-03 | 2.57E-02 |
| CD53 | 0.929 | 9.639 | 3.953 | 9.96E-05 | 1.13E-03 |
| PSMB10 | 0.927 | 9.059 | 5.831 | 1.05E-07 | 2.71E-06 |
| C2 | 0.927 | 8.818 | 3.342 | 7.06E-04 | 6.24E-03 |
| DDIT3 | 0.926 | 8.847 | 3.141 | 1.29E-03 | 1.04E-02 |
| ZEB2 | 0.926 | 8.916 | 4.334 | 2.71E-05 | 3.40E-04 |
| LAMA5 | 0.924 | 9.451 | 3.138 | 1.30E-03 | 1.05E-02 |
| ATP2A2 | 0.923 | 9.483 | 5.114 | 1.62E-06 | 2.85E-05 |
| LCP1 | 0.919 | 9.461 | 3.604 | 3.11E-04 | 3.08E-03 |
| SIRPA | 0.910 | 9.089 | 5.091 | 1.76E-06 | 3.06E-05 |
| ITGA5 | 0.900 | 9.093 | 3.507 | 4.22E-04 | 4.03E-03 |
| RHOA | 0.897 | 9.787 | 3.646 | 2.72E-04 | 2.77E-03 |
| HK1 | 0.892 | 9.384 | 6.681 | 3.74E-09 | 1.31E-07 |
| TGFBR2 | 0.883 | 10.060 | 3.387 | 6.13E-04 | 5.57E-03 |
| BRD4 | 0.881 | 9.493 | 4.735 | 6.51E-06 | 9.42E-05 |
| LTBP1 | 0.875 | 8.716 | 2.736 | 4.04E-03 | 2.80E-02 |
| PEBP1 | 0.872 | 9.606 | 2.805 | 3.35E-03 | 2.38E-02 |
| U2AF1 | 0.871 | 9.414 | 4.423 | 1.99E-05 | 2.57E-04 |
| NDUFA1 | 0.862 | 9.731 | 3.599 | 3.16E-04 | 3.11E-03 |
| C4A-B | 0.860 | 9.518 | 2.502 | 7.51E-03 | 4.68E-02 |
| FLNB | 0.860 | 9.313 | 3.668 | 2.54E-04 | 2.62E-03 |
| FUBP1 | 0.856 | 9.347 | 5.016 | 2.33E-06 | 3.86E-05 |
| CCL15 | 0.855 | 9.385 | 4.396 | 2.18E-05 | 2.77E-04 |
| PARP9 | 0.852 | 9.343 | 4.921 | 3.31E-06 | 5.29E-05 |
| PRKACA | 0.851 | 8.852 | 4.797 | 5.20E-06 | 7.76E-05 |
| DTX3L | 0.846 | 9.100 | 5.046 | 2.08E-06 | 3.57E-05 |
| SLC25A1 | 0.832 | 9.134 | 3.812 | 1.59E-04 | 1.71E-03 |
| LGALS9 | 0.832 | 9.140 | 3.185 | 1.13E-03 | 9.35E-03 |
| SMARCA4 | 0.831 | 9.063 | 5.027 | 2.24E-06 | 3.80E-05 |
| JUP | 0.826 | 9.245 | 3.102 | 1.44E-03 | 1.15E-02 |
| IGF2R | 0.826 | 9.243 | 4.152 | 5.09E-05 | 6.01E-04 |
| DST | 0.826 | 8.718 | 3.365 | 6.58E-04 | 5.91E-03 |
| OAT | 0.826 | 8.929 | 4.741 | 6.37E-06 | 9.30E-05 |
| CSF3R | 0.824 | 8.898 | 2.734 | 4.07E-03 | 2.81E-02 |
| TAP2 | 0.823 | 9.623 | 2.847 | 2.98E-03 | 2.19E-02 |
| ATP5ME | 0.821 | 9.880 | 3.857 | 1.37E-04 | 1.50E-03 |
| PTEN | 0.820 | 8.906 | 3.899 | 1.19E-04 | 1.32E-03 |
| SHC1 | 0.817 | 9.448 | 3.201 | 1.08E-03 | 9.00E-03 |
| IDH2 | 0.816 | 9.175 | 3.729 | 2.08E-04 | 2.19E-03 |
| SGK1 | 0.804 | 9.027 | 2.572 | 6.25E-03 | 4.05E-02 |
| C1QBP | 0.803 | 9.061 | 3.829 | 1.51E-04 | 1.64E-03 |
| ATOX1 | 0.800 | 9.071 | 5.632 | 2.27E-07 | 5.25E-06 |
| GPX4 | 0.791 | 9.956 | 2.815 | 3.26E-03 | 2.34E-02 |
| MAPKAPK2 | 0.788 | 9.282 | 3.667 | 2.54E-04 | 2.62E-03 |
| MAPK3 | 0.786 | 8.784 | 4.598 | 1.07E-05 | 1.51E-04 |
| TYK2 | 0.785 | 9.072 | 4.890 | 3.71E-06 | 5.83E-05 |
| FCGR2A | 0.782 | 8.870 | 3.346 | 6.97E-04 | 6.21E-03 |
| EIF4EBP1 | 0.779 | 8.822 | 3.148 | 1.26E-03 | 1.02E-02 |
| RAD21 | 0.775 | 9.198 | 3.264 | 8.92E-04 | 7.55E-03 |
| PRKCD | 0.772 | 8.887 | 4.869 | 4.01E-06 | 6.19E-05 |
| MS4A6A | 0.771 | 9.283 | 2.659 | 4.97E-03 | 3.34E-02 |
| UBA7 | 0.770 | 8.859 | 4.542 | 1.30E-05 | 1.79E-04 |
| GNG12 | 0.765 | 9.043 | 3.341 | 7.07E-04 | 6.24E-03 |
| TRAF7 | 0.764 | 8.947 | 4.551 | 1.26E-05 | 1.75E-04 |
| PML | 0.756 | 9.005 | 3.018 | 1.84E-03 | 1.44E-02 |
| PSMB5 | 0.755 | 9.193 | 3.698 | 2.31E-04 | 2.40E-03 |
| ITPK1 | 0.749 | 8.880 | 3.446 | 5.12E-04 | 4.79E-03 |
| PSMB8 | 0.744 | 9.238 | 3.178 | 1.15E-03 | 9.49E-03 |
| API5 | 0.737 | 8.811 | 4.508 | 1.47E-05 | 1.97E-04 |
| IFI16 | 0.737 | 8.657 | 2.488 | 7.77E-03 | 4.78E-02 |
| BCL6 | 0.737 | 9.083 | 3.242 | 9.53E-04 | 8.02E-03 |
| CFI | 0.733 | 8.797 | 2.547 | 6.68E-03 | 4.25E-02 |
| ABL1 | 0.726 | 8.798 | 3.328 | 7.36E-04 | 6.38E-03 |
| CSF1 | 0.719 | 8.808 | 2.697 | 4.50E-03 | 3.07E-02 |
| NDUFB10 | 0.709 | 9.215 | 3.372 | 6.43E-04 | 5.81E-03 |
| PARP4 | 0.706 | 8.960 | 3.616 | 3.00E-04 | 2.98E-03 |
| PFKFB3 | 0.704 | 9.115 | 2.556 | 6.52E-03 | 4.18E-02 |
| GLS | 0.703 | 8.825 | 2.732 | 4.09E-03 | 2.81E-02 |
| NFE2L2 | 0.699 | 9.332 | 3.314 | 7.67E-04 | 6.58E-03 |
| PSMB7 | 0.695 | 9.198 | 3.703 | 2.27E-04 | 2.37E-03 |
| FURIN | 0.691 | 9.272 | 2.623 | 5.47E-03 | 3.61E-02 |
| SIGLEC1 | 0.684 | 8.758 | 3.114 | 1.39E-03 | 1.12E-02 |
| SERINC1 | 0.682 | 9.016 | 2.577 | 6.18E-03 | 4.02E-02 |
| TWF1 | 0.681 | 8.924 | 4.175 | 4.71E-05 | 5.59E-04 |
| IFI35 | 0.680 | 9.026 | 2.490 | 7.73E-03 | 4.78E-02 |
| IFNGR2 | 0.679 | 9.439 | 3.327 | 7.38E-04 | 6.38E-03 |
| DDB2 | 0.676 | 8.421 | 3.106 | 1.43E-03 | 1.14E-02 |
| PCNA | 0.674 | 8.884 | 2.474 | 8.05E-03 | 4.90E-02 |
| IRF9 | 0.668 | 9.101 | 4.300 | 3.06E-05 | 3.73E-04 |
| NOTCH1 | 0.668 | 8.847 | 2.967 | 2.13E-03 | 1.61E-02 |
| LTBR | 0.665 | 9.150 | 2.983 | 2.03E-03 | 1.55E-02 |
| SMAD4 | 0.653 | 8.630 | 3.172 | 1.17E-03 | 9.62E-03 |
| PCK2 | 0.652 | 8.514 | 2.953 | 2.22E-03 | 1.67E-02 |
| ETS1 | 0.648 | 8.905 | 2.579 | 6.13E-03 | 4.01E-02 |
| FH | 0.646 | 8.682 | 3.545 | 3.76E-04 | 3.61E-03 |
| IL13RA1 | 0.645 | 9.073 | 2.743 | 3.97E-03 | 2.77E-02 |
| BAGE | 0.640 | 9.522 | 2.821 | 3.21E-03 | 2.31E-02 |
| RELA | 0.638 | 9.181 | 3.658 | 2.62E-04 | 2.68E-03 |
| CUL1 | 0.637 | 8.791 | 4.010 | 8.25E-05 | 9.49E-04 |
| SF3A1 | 0.636 | 8.829 | 3.395 | 6.00E-04 | 5.47E-03 |
| MBNL1 | 0.632 | 9.157 | 2.528 | 7.01E-03 | 4.41E-02 |
| COX4I1 | 0.624 | 9.564 | 2.838 | 3.05E-03 | 2.23E-02 |
| GNA11 | 0.624 | 9.045 | 3.014 | 1.86E-03 | 1.45E-02 |
| E2F4 | 0.615 | 8.931 | 2.993 | 1.98E-03 | 1.52E-02 |
| DNMT1 | 0.602 | 8.719 | 3.302 | 7.95E-04 | 6.76E-03 |
| CDK4 | 0.599 | 8.653 | 2.603 | 5.76E-03 | 3.79E-02 |
| KMT2D | 0.599 | 8.883 | 3.162 | 1.21E-03 | 9.87E-03 |
| SPOP | 0.597 | 8.754 | 2.832 | 3.11E-03 | 2.26E-02 |
| SERINC5 | 0.585 | 8.560 | 2.534 | 6.90E-03 | 4.38E-02 |
| MAPK14 | 0.584 | 8.756 | 3.084 | 1.52E-03 | 1.20E-02 |
| PTPN11 | 0.578 | 9.125 | 2.652 | 5.06E-03 | 3.38E-02 |
| PYCR2 | 0.576 | 8.510 | 2.822 | 3.20E-03 | 2.31E-02 |
| CKLF | 0.575 | 8.820 | 2.811 | 3.29E-03 | 2.35E-02 |
| ARID1A | 0.570 | 8.545 | 2.874 | 2.77E-03 | 2.04E-02 |
| TNFRSF14 | 0.559 | 8.985 | 3.397 | 5.96E-04 | 5.47E-03 |
| UQCR11 | 0.558 | 8.619 | 2.951 | 2.23E-03 | 1.67E-02 |
| SF3A3 | 0.554 | 8.864 | 2.671 | 4.82E-03 | 3.26E-02 |
| NASP | 0.530 | 8.950 | 2.484 | 7.86E-03 | 4.82E-02 |
| NDUFS7 | 0.525 | 8.851 | 2.530 | 6.98E-03 | 4.41E-02 |
| RBX1 | 0.515 | 8.778 | 2.768 | 3.71E-03 | 2.60E-02 |

*Highlighted genes (*LY6E, IFI27* & *IFI6)* also found to be upregulated in SARS-CoV-2 patients when compared to pH1N1 influenza samples.

**Extended Data Table 8.** **Genes differentially expressed in the lungs of SARS-CoV-2 patients with high viral load compared with patients with low viral load (Region based analysis).**

| **Gene name** | **Fold Change (log2)** | **Average Expression** | **t-statistics** | **p-value** | **Adjusted p-value** |
| --- | --- | --- | --- | --- | --- |
| ***S*** | 2.415 | 8.900 | 5.616 | 4.985E-07 | 2.350E-04 |
| ***ORF1ab*** | 2.365 | 9.036 | 5.611 | 5.076E-07 | 2.350E-04 |
| ***CXCL11*** | 0.946 | 8.291 | 4.533 | 2.738E-05 | 2.414E-03 |
| ***RSAD2*** | 0.669 | 8.662 | 3.014 | 3.737E-03 | 4.524E-02 |
| ***IL1RN*** | 0.584 | 8.130 | 4.057 | 1.421E-04 | 6.923E-03 |
| ***ANGPTL4*** | 0.555 | 8.008 | 3.216 | 2.073E-03 | 3.392E-02 |
| ***CD274*** | 0.531 | 8.076 | 6.292 | 3.621E-08 | 6.706E-05 |
| ***CCL8*** | 0.466 | 7.679 | 5.045 | 4.276E-06 | 1.320E-03 |
| ***IL15RA*** | 0.437 | 8.696 | 3.277 | 1.726E-03 | 3.043E-02 |
| ***SOCS1*** | 0.431 | 8.547 | 3.881 | 2.560E-04 | 9.676E-03 |
| ***CCL7*** | 0.401 | 7.816 | 4.008 | 1.679E-04 | 7.585E-03 |
| ***OASL*** | 0.382 | 8.196 | 2.989 | 4.020E-03 | 4.653E-02 |
| ***SARS-CoV-2-Neg*** | 0.357 | 7.714 | 4.643 | 1.845E-05 | 2.136E-03 |
| ***IL19*** | 0.305 | 7.750 | 3.648 | 5.454E-04 | 1.554E-02 |
| ***CSF2RB*** | 0.304 | 8.473 | 3.213 | 2.088E-03 | 3.392E-02 |
| ***CALML5*** | 0.296 | 7.632 | 3.022 | 3.655E-03 | 4.498E-02 |
| ***WNT2*** | 0.293 | 7.858 | 3.429 | 1.087E-03 | 2.314E-02 |
| ***FGF17*** | 0.290 | 7.318 | 3.552 | 7.402E-04 | 1.828E-02 |
| ***ACE2*** | 0.279 | 7.833 | 3.577 | 6.839E-04 | 1.759E-02 |
| ***TLR8*** | 0.277 | 8.246 | 3.190 | 2.238E-03 | 3.508E-02 |
| ***CLECL1*** | 0.269 | 7.774 | 3.056 | 3.312E-03 | 4.303E-02 |
| ***MASP2*** | 0.244 | 7.920 | 3.034 | 3.527E-03 | 4.414E-02 |
| ***PPP3R2*** | 0.241 | 7.704 | 3.107 | 2.861E-03 | 3.988E-02 |
| ***RIMKLA*** | 0.237 | 7.887 | 3.913 | 2.302E-04 | 9.546E-03 |
| ***MAGEB2*** | 0.227 | 7.673 | 3.039 | 3.479E-03 | 4.383E-02 |
| ***RET*** | 0.220 | 8.077 | 3.400 | 1.186E-03 | 2.392E-02 |
| ***OTC*** | 0.218 | 7.887 | 3.055 | 3.322E-03 | 4.303E-02 |
| ***OTOA*** | 0.216 | 7.789 | 3.100 | 2.915E-03 | 3.988E-02 |
| ***SH2B2*** | 0.207 | 8.230 | 3.101 | 2.905E-03 | 3.988E-02 |
| ***MAPK8IP2*** | 0.206 | 8.026 | 3.226 | 2.012E-03 | 3.357E-02 |
| ***CCR3*** | 0.201 | 7.703 | 2.971 | 4.222E-03 | 4.827E-02 |
| ***IKBKE*** | 0.197 | 8.207 | 3.117 | 2.774E-03 | 3.988E-02 |
| ***MUC2*** | 0.184 | 8.239 | 3.010 | 3.781E-03 | 4.548E-02 |
| ***STAT5B*** | -0.169 | 8.393 | -3.147 | 2.544E-03 | 3.831E-02 |
| ***POLR2D*** | -0.195 | 8.264 | -3.447 | 1.027E-03 | 2.239E-02 |
| ***PIAS4*** | -0.196 | 8.294 | -3.510 | 8.437E-04 | 1.930E-02 |
| ***APC*** | -0.201 | 7.947 | -3.306 | 1.581E-03 | 2.892E-02 |
| ***PPP3CA*** | -0.208 | 8.486 | -3.020 | 3.673E-03 | 4.498E-02 |
| ***ITCH*** | -0.209 | 8.672 | -3.158 | 2.460E-03 | 3.735E-02 |
| ***ERCC3*** | -0.213 | 8.212 | -3.049 | 3.379E-03 | 4.345E-02 |
| ***IFNAR1*** | -0.213 | 8.297 | -3.639 | 5.615E-04 | 1.575E-02 |
| ***UQCR11*** | -0.214 | 8.619 | -3.180 | 2.306E-03 | 3.558E-02 |
| ***SF3A3*** | -0.218 | 8.864 | -2.963 | 4.329E-03 | 4.888E-02 |
| ***API5*** | -0.224 | 8.811 | -3.000 | 3.889E-03 | 4.597E-02 |
| ***PSMD7*** | -0.225 | 8.980 | -3.162 | 2.431E-03 | 3.721E-02 |
| ***TPR*** | -0.229 | 8.771 | -3.120 | 2.752E-03 | 3.988E-02 |
| ***DNMT1*** | -0.230 | 8.719 | -3.424 | 1.102E-03 | 2.319E-02 |
| ***RAF1*** | -0.232 | 8.798 | -2.997 | 3.922E-03 | 4.597E-02 |
| ***RIN1*** | -0.232 | 8.270 | -3.313 | 1.549E-03 | 2.892E-02 |
| ***CKLF*** | -0.237 | 8.820 | -3.135 | 2.632E-03 | 3.899E-02 |
| ***SRP54*** | -0.237 | 8.832 | -2.975 | 4.177E-03 | 4.805E-02 |
| ***KMT2D*** | -0.238 | 8.883 | -3.317 | 1.531E-03 | 2.892E-02 |
| ***HMGB1*** | -0.238 | 8.644 | -3.096 | 2.950E-03 | 3.988E-02 |
| ***MAPK1*** | -0.244 | 8.648 | -3.229 | 1.996E-03 | 3.357E-02 |
| ***PVRIG*** | -0.245 | 8.911 | -3.006 | 3.824E-03 | 4.569E-02 |
| ***TGFB3*** | -0.247 | 8.411 | -2.997 | 3.922E-03 | 4.597E-02 |
| ***KMT2C*** | -0.249 | 8.808 | -3.543 | 7.620E-04 | 1.857E-02 |
| ***FEN1*** | -0.250 | 8.094 | -3.416 | 1.129E-03 | 2.349E-02 |
| ***TBL1XR1*** | -0.252 | 9.164 | -3.105 | 2.875E-03 | 3.988E-02 |
| ***NDUFA2*** | -0.253 | 9.007 | -3.307 | 1.576E-03 | 2.892E-02 |
| ***ROCK1*** | -0.254 | 8.777 | -3.739 | 4.073E-04 | 1.245E-02 |
| ***HDAC2*** | -0.257 | 8.493 | -3.107 | 2.858E-03 | 3.988E-02 |
| ***ATRX*** | -0.260 | 8.724 | -3.835 | 2.979E-04 | 1.012E-02 |
| ***KRT10*** | -0.262 | 8.395 | -3.296 | 1.631E-03 | 2.905E-02 |
| ***TXN2*** | -0.263 | 8.965 | -3.385 | 1.244E-03 | 2.477E-02 |
| ***GOT2*** | -0.263 | 8.994 | -3.408 | 1.157E-03 | 2.381E-02 |
| ***TGFBR1*** | -0.267 | 8.665 | -3.784 | 3.518E-04 | 1.143E-02 |
| ***IRF3*** | -0.273 | 8.749 | -3.340 | 1.427E-03 | 2.781E-02 |
| ***FH*** | -0.273 | 8.682 | -3.737 | 4.100E-04 | 1.245E-02 |
| ***CDKN2C*** | -0.274 | 7.931 | -3.200 | 2.175E-03 | 3.502E-02 |
| ***PALMD*** | -0.275 | 8.157 | -3.274 | 1.742E-03 | 3.043E-02 |
| ***NDUFA3*** | -0.283 | 9.207 | -3.088 | 3.023E-03 | 4.011E-02 |
| ***SMARCB1*** | -0.286 | 8.735 | -4.142 | 1.068E-04 | 5.650E-03 |
| ***ATF2*** | -0.288 | 8.348 | -4.294 | 6.329E-05 | 4.042E-03 |
| ***PARP4*** | -0.289 | 8.960 | -3.380 | 1.262E-03 | 2.486E-02 |
| ***COX4I1*** | -0.291 | 9.564 | -3.430 | 1.082E-03 | 2.314E-02 |
| ***PTPN11*** | -0.291 | 9.125 | -3.770 | 3.686E-04 | 1.177E-02 |
| ***NOTCH1*** | -0.292 | 8.847 | -3.040 | 3.470E-03 | 4.383E-02 |
| ***FUBP1*** | -0.296 | 9.347 | -3.512 | 8.382E-04 | 1.930E-02 |
| ***LRP6*** | -0.298 | 8.132 | -3.477 | 9.349E-04 | 2.086E-02 |
| ***SMAD4*** | -0.298 | 8.630 | -3.694 | 4.710E-04 | 1.363E-02 |
| ***DAXX*** | -0.298 | 8.341 | -4.749 | 1.264E-05 | 1.672E-03 |
| ***NFE2L2*** | -0.299 | 9.332 | -3.304 | 1.593E-03 | 2.892E-02 |
| ***CREBBP*** | -0.301 | 8.512 | -4.209 | 8.489E-05 | 5.240E-03 |
| ***NDUFB1*** | -0.301 | 9.165 | -4.336 | 5.465E-05 | 3.771E-03 |
| ***TRAF7*** | -0.306 | 8.947 | -4.108 | 1.199E-04 | 6.167E-03 |
| ***SPOP*** | -0.306 | 8.754 | -3.973 | 1.886E-04 | 8.317E-03 |
| ***JAK1*** | -0.307 | 9.550 | -3.087 | 3.032E-03 | 4.011E-02 |
| ***MAVS*** | -0.307 | 8.688 | -3.491 | 8.948E-04 | 2.021E-02 |
| ***PDGFA*** | -0.310 | 8.602 | -3.089 | 3.013E-03 | 4.011E-02 |
| ***ATP2A2*** | -0.317 | 9.483 | -3.327 | 1.485E-03 | 2.835E-02 |
| ***PRKACA*** | -0.321 | 8.852 | -3.602 | 6.317E-04 | 1.696E-02 |
| ***BRD2*** | -0.324 | 9.546 | -2.991 | 3.995E-03 | 4.653E-02 |
| ***H2AX*** | -0.329 | 8.721 | -3.752 | 3.905E-04 | 1.226E-02 |
| ***MCM7*** | -0.333 | 8.619 | -3.869 | 2.664E-04 | 9.866E-03 |
| ***LEPR*** | -0.334 | 8.243 | -3.571 | 6.969E-04 | 1.768E-02 |
| ***PSMB5*** | -0.337 | 9.193 | -3.719 | 4.338E-04 | 1.296E-02 |
| ***ABL1*** | -0.339 | 8.798 | -3.534 | 7.819E-04 | 1.866E-02 |
| ***TP53*** | -0.340 | 8.576 | -4.612 | 2.063E-05 | 2.189E-03 |
| ***PRKDC*** | -0.344 | 8.734 | -4.303 | 6.126E-05 | 4.042E-03 |
| ***CDKN1B*** | -0.346 | 9.159 | -3.533 | 7.860E-04 | 1.866E-02 |
| ***ITGA6*** | -0.348 | 8.091 | -3.142 | 2.579E-03 | 3.853E-02 |
| ***PPP3R1*** | -0.357 | 8.489 | -4.851 | 8.717E-06 | 1.535E-03 |
| ***COX6B1*** | -0.362 | 9.843 | -3.018 | 3.692E-03 | 4.498E-02 |
| ***OAT*** | -0.375 | 8.929 | -4.838 | 9.146E-06 | 1.535E-03 |
| ***P4HA2*** | -0.380 | 8.858 | -3.271 | 1.759E-03 | 3.045E-02 |
| ***STMN1*** | -0.385 | 8.393 | -3.891 | 2.474E-04 | 9.546E-03 |
| ***CD99*** | -0.393 | 9.679 | -3.577 | 6.827E-04 | 1.759E-02 |
| ***ITGA1*** | -0.396 | 8.799 | -3.466 | 9.671E-04 | 2.132E-02 |
| ***SERINC3*** | -0.404 | 9.130 | -4.568 | 2.418E-05 | 2.239E-03 |
| ***DST*** | -0.405 | 8.718 | -3.336 | 1.445E-03 | 2.788E-02 |
| ***THBD*** | -0.406 | 9.088 | -3.019 | 3.685E-03 | 4.498E-02 |
| ***NDUFA11*** | -0.408 | 9.594 | -3.833 | 2.996E-04 | 1.012E-02 |
| ***MKI67*** | -0.409 | 8.237 | -3.298 | 1.620E-03 | 2.905E-02 |
| ***ITGAV*** | -0.415 | 9.074 | -3.188 | 2.254E-03 | 3.508E-02 |
| ***NDUFS8*** | -0.415 | 8.971 | -4.370 | 4.856E-05 | 3.597E-03 |
| ***RAD21*** | -0.417 | 9.198 | -3.918 | 2.262E-04 | 9.546E-03 |
| ***NDUFA1*** | -0.417 | 9.731 | -3.620 | 5.967E-04 | 1.625E-02 |
| ***CDK4*** | -0.419 | 8.653 | -5.278 | 1.793E-06 | 6.642E-04 |
| ***SF3B1*** | -0.427 | 9.651 | -3.799 | 3.353E-04 | 1.109E-02 |
| ***RHOA*** | -0.439 | 9.787 | -3.561 | 7.194E-04 | 1.801E-02 |
| ***RANBP2*** | -0.441 | 9.123 | -4.815 | 9.945E-06 | 1.535E-03 |
| ***TYMS*** | -0.447 | 8.301 | -3.116 | 2.785E-03 | 3.988E-02 |
| ***THY1*** | -0.451 | 8.833 | -3.122 | 2.736E-03 | 3.988E-02 |
| ***PEBP1*** | -0.465 | 9.606 | -3.042 | 3.453E-03 | 4.383E-02 |
| ***CCND2*** | -0.466 | 8.729 | -3.839 | 2.935E-04 | 1.012E-02 |
| ***PDGFRB*** | -0.466 | 9.320 | -2.954 | 4.442E-03 | 4.985E-02 |
| ***ANP32B*** | -0.469 | 9.170 | -4.150 | 1.039E-04 | 5.650E-03 |
| ***LAMA5*** | -0.473 | 9.451 | -3.097 | 2.944E-03 | 3.988E-02 |
| ***TECR*** | -0.475 | 9.326 | -4.975 | 5.523E-06 | 1.461E-03 |
| ***ITGAX*** | -0.481 | 9.025 | -3.080 | 3.094E-03 | 4.064E-02 |
| ***C4A-B*** | -0.484 | 9.518 | -2.969 | 4.251E-03 | 4.830E-02 |
| ***ST6GAL1*** | -0.498 | 8.508 | -4.435 | 3.872E-05 | 2.988E-03 |
| ***SPRY1*** | -0.504 | 8.263 | -4.574 | 2.367E-05 | 2.239E-03 |
| ***H3-3A*** | -0.515 | 10.042 | -4.083 | 1.303E-04 | 6.523E-03 |
| ***PTEN*** | -0.518 | 8.906 | -5.953 | 1.362E-07 | 1.261E-04 |
| ***TPM4*** | -0.533 | 10.618 | -3.101 | 2.904E-03 | 3.988E-02 |
| ***SLC1A5*** | -0.534 | 9.260 | -4.012 | 1.652E-04 | 7.585E-03 |
| ***APP*** | -0.536 | 10.607 | -3.189 | 2.247E-03 | 3.508E-02 |
| ***PRDX5*** | -0.544 | 9.811 | -3.624 | 5.894E-04 | 1.625E-02 |
| ***TUBB*** | -0.558 | 9.530 | -4.185 | 9.207E-05 | 5.501E-03 |
| ***HSP90B1*** | -0.569 | 10.113 | -3.219 | 2.054E-03 | 3.392E-02 |
| ***NOTCH3*** | -0.576 | 8.919 | -4.749 | 1.261E-05 | 1.672E-03 |
| ***CTNNB1*** | -0.581 | 9.983 | -4.456 | 3.585E-05 | 2.886E-03 |
| ***FLNC*** | -0.583 | 8.103 | -3.906 | 2.354E-04 | 9.546E-03 |
| ***RPS27A*** | -0.615 | 10.665 | -3.400 | 1.188E-03 | 2.392E-02 |
| ***RPL7A*** | -0.623 | 11.626 | -3.227 | 2.007E-03 | 3.357E-02 |
| ***SREBF1*** | -0.655 | 9.546 | -3.583 | 6.709E-04 | 1.759E-02 |
| ***F13A1*** | -0.672 | 8.789 | -3.190 | 2.240E-03 | 3.508E-02 |
| ***LDHB*** | -0.679 | 9.866 | -3.855 | 2.791E-04 | 1.012E-02 |
| ***CD9*** | -0.725 | 10.245 | -3.510 | 8.442E-04 | 1.930E-02 |
| ***MCAM*** | -0.733 | 8.993 | -4.464 | 3.495E-05 | 2.886E-03 |
| ***ITGB1*** | -0.748 | 10.627 | -4.604 | 2.128E-05 | 2.189E-03 |
| ***GNAS*** | -0.751 | 11.510 | -4.154 | 1.025E-04 | 5.650E-03 |
| ***FLNA*** | -0.766 | 10.894 | -3.266 | 1.782E-03 | 3.055E-02 |
| ***RHOB*** | -0.785 | 10.349 | -3.695 | 4.698E-04 | 1.363E-02 |
| ***H3C8*** | -0.788 | 8.504 | -4.706 | 1.476E-05 | 1.822E-03 |
| ***RPS6*** | -0.831 | 11.502 | -3.900 | 2.401E-04 | 9.546E-03 |
| ***RPL23*** | -0.852 | 11.713 | -4.159 | 1.005E-04 | 5.650E-03 |
| ***H3C10*** | -0.873 | 10.187 | -4.047 | 1.469E-04 | 6.975E-03 |
| ***TPM1*** | -0.902 | 9.776 | -4.869 | 8.144E-06 | 1.535E-03 |
| ***SFRP2*** | -0.946 | 8.442 | -3.895 | 2.442E-04 | 9.546E-03 |
| ***H3C2*** | -0.998 | 8.791 | -4.823 | 9.631E-06 | 1.535E-03 |
| ***ACTA2*** | -1.103 | 9.385 | -4.334 | 5.498E-05 | 3.771E-03 |
| ***C3*** | -1.110 | 10.763 | -3.832 | 3.006E-04 | 1.012E-02 |

Genes upregulated genes in high viral load (green) compared with low vial load SARS-CoV-2 lung samples. Downregulated genes are shown in red.
